# IP10 levels refine severity prognostication in COVID-19 and implicate causes of clinical deterioration: lessons for pandemic preparedness

**DOI:** 10.1101/2024.06.15.24308935

**Authors:** Abhishek Das, Jia Wei, Duncan McKenzie, Luke Snell, Shruthi Sasidharan, Pierre Vantourout, Iva Zlatareva, Blair Merrick, Benjamin Thomas, Vasista Adiga, Khiyam Hussain, Rahul Batra, Daniel Davies, Jia Su, Karen Bisnauthsing, Lauren Martinez, Asma Ahmed, Hima Bindhu, Nirutha Chetan, Maria Macrina, Himanshu Tripathi, Geraldine O’Hara, Cassandra Fairhead, Gabija Drazdauskaite, Helen Lock, Mary Dias, Mohammad A. A. Ibrahim, Thomas Hayday, George D’Souza, Jonathan Edgeworth, Annapurna Vyakarnam, Koen B Pouwels, Adrian C. Hayday

## Abstract

**Background:** Emerging pandemics place immense strains on healthcare systems that may be ameliorated by rapid development of biomarkers whose measurements may predict disease severity and additionally inform about disease causation. Conspicuously, such routine measures rarely include immunological cytokines or chemokines, despite their contributions to host protection and immunopathology.

**Methods:** Multiplex bead-array and ELISA-based serum cytokine and chemokine measurements, routinely employed clinical laboratory measures, and clinical outcomes were collectively fed into predictive model development for prognostication of COVID-19 severity in an unvaccinated UK cohort (Discovery; early-to-mid 2020), with subsequent external validation among a pauci-vaccinated UK cohort (early 2021) and part-vaccinated India cohort (early 2022 to early 2023). Correlates of disease severity were assessed by high-content spectral flow cytometry.

**Findings:** Incorporating a practical test for the chemokine IP10 (a.k.a. CXCL10) alongside routine clinical laboratory assays increased at-admission test accuracy to prognosticate intensive care requirements or in-hospital mortality at 30 days in the Discovery and Validation cohorts. In the India cohort, high IP10 levels predicted terminal deterioration among unvaccinated persons. High-resolution immune-profiling within subsets of the Discovery and India cohorts associated a T cell-centric signature with disease severity and with high IP10 levels, thereby identifying candidate drivers of COVID-19 deterioration.

**Conclusions:** IP10 levels measured at or around hospital admission offer a practical biomarker enhancing COVID-19 patient outcome prognostication, particluarly in unvaccinated individuals, and offer mechanistic insights into pathogenesis. Thus, prompt application of systems immune-profiling in future pandemics might rapidly identify prognostic and mechanistic biomarkers of patient deterioration, aiding clinical decision-making at a time of severe healthcare strain.

**Funding:** Medical Research Council grant, CARDINNATE.

## INTRODUCTION

As part of pandemic preparedness, some compelling lessons should clearly be learned from COVID-19^1^. Among them, clinical decision-making might be more confident and demands on healthcare resources ameliorated, by early identification of practical biomarkers that at hospital admission can discriminate those patients most likely to deteriorate^2,3^. Such biomarkers might support early triage to intensive care or targeted therapy, potentially mitigating against severe disease and / or long-term health sequelae. Linked to this, the nature of the relevant biomarker(s) might expose mechanistic contributors to short-term and / or long-term disease severity, thereby improving our understanding and aiding the development of therapeutic strategies.

Given the central role of the immune system in mediating anti-viral protection and / or pathology, cytokines and chemokines might seem strong candidates for such biomarkers^4–8^. However, these are rarely included among the panels of easily measured clinical markers, e.g., C-reactive protein (CRP), albumin, and neutrophil counts, composed for various infectious and inflammatory disease settings. Indeed, whereas the COVID-19 pandemic witnessed the rapid development of several prognostic scores with adequate predictive performance, including the ISARIC mortality and deterioration scores^2,9–11^, the potential utility of cytokine or chemokine measurements was commonly overlooked.

In fact, our own immune-profiling of hospitalized COVID-19 patients in the first pandemic wave highlighted that patients were more likely to deteriorate if, at or around the time of hospital admission, they displayed high serum concentrations of a triad of analytes, the chemokine IP10 (a.k.a. CXCL10) and two cytokines, IL-10 and IL-6^4^. Now we report the validation of this prognostic potential in a second wave of the pandemic in the same hospitals but caused mostly by a different SARS-CoV-2 strain. Thus, our data provide the rationale and practical means to incorporate IP10 as a cardinal biomarker associated with underpinning immunopathology, to facilitate early detection of patients at risk of subsequent deterioration.

Additionally, our study addresses an invariable aspect of pandemics, namely that the phenotype of COVID19-associated hospitalization has rapidly transitioned, driven by a combination of shifts in viral evolution, host-directed immunotherapies, and vaccination. Indeed, today’s COVID-19 disease cohorts are very different to those in the pre-vaccine era. Nevertheless, we hypothesized that if high IP10 levels reflect core traits potentially contributing to host pathology, then those may be preserved in patients who progress to organ failure and death despite improved therapies being available. To test this, we examined an highly heterogeneous cohort of SARS-CoV-2-infected patients hospitalized in 2022-2023 in Bangalore, specifically demonstrating that IP10 measurements made at or around the time of hospital admission continued to relate to the most extreme downstream outcomes.

Finally, we have asked whether the correlation of at-admission high IP10 levels with deterioration and/or death across the three cohorts might provide insights into cellular immune factors likely contributing to pathology. To this end, we employed unbiased, high-resolution, high-content immune-profiling that has strongly implicated early T cell dysregulation. Collectively, these findings support the prompt application of systems immune-profiling in future pandemics in order to rapidly identify prognostic and mechanistic biomarkers of patient deterioration.

## STAR METHODS

### Study population

Our study focussed on 659 patients.

First, 454 patients with PCR-confirmed SARS-CoV-2 were recruited from a UK hospital for the Discovery and Validation cohorts. Residual serum samples, collected at the point of hospital admission (n=426 (94%) on day 0 or 1; n=28 (6%) on day 2) for routine biochemistry analysis were retrieved, aliquoted, pseudoanonymised and stored at −80°C for later analysis. All recruitment was prospective, with the exception of eight COVID-19 serum samples derived from our initial study^4^, for whom a sample was available from between day 0-2 following admission. Likewise, where comparisons were made to healthy controls, recruitment of this population leveraged a mixture of previously published serum cytokine/chemokine data (n=55) generated during our initial study, and prospectively collected healthy donor samples (n=6).

Second, for the India cohort, we prospectively recruited 88 patients with PCR-confirmed SARS-CoV-2 and 100 patients without current or prior documented history of COVID-19, from a tertiary hospital in Bangalore. Samples were collected between January 2022 and February 2023, at a time when most documented cases were caused by the Omicron variant, although sequencing was not performed. Serum was available from all patients, whilst stored, high quality peripheral blood mononuclear cells (PBMC) for cellular immunophenotyping were available from only a sub-cohort (SARS-CoV-2^+^, n=34 and patients without COVID-19, n=51). In order to compare cellular traits against a UK cohort in whom a similar proportion of patients deteriorated, we additionally retrieved PBMC samples from 17 UK patients with COVID-19 from the first 2020 wave.

Use of residual or discarded serum from routine clinical samples was granted ethical approval through the Ethical Determination In Coronavirus diagnosTic developmentS (EDICTS) study: REC Approval: 20/SC/0310, by the South Central Hampshire B Research Ethics committee in 2020. A subset of clinical information was collected by the direct care team and provided to researchers in a linked-anonymised fashion. A standard operating procedure was in place (IRAS Project ID:237180) to ensure that the material removed for the study did not have an adverse effect on the routine clinical diagnostics undertaken on that sample. A further 8 patients with COVID-19 and all healthy donors were recruited following informed consent and the study protocol for patient recruitment and sampling was approved by the committee of the Infectious Diseases Biobank of King’s College London with reference number COV-250320 (patients) and MJ1-031218b (healthy donors). Both approvals were granted under the terms of the Infectious Disease Biobank’s ethics permission (reference 19/SC/0232) granted by the South Central Hampshire B Research Ethics Committee in 2019. Patient and control samples and data were anonymized at the point of sample collection by research nursing staff or clinicians involved in the COVID-IP project^4^.

Collection of blood samples for the study entitled ‘Variation in innate immune activation and cardiovascular disease risk as drivers of COVID-19 outcome in South Asians in UK and India’ was approved by the Institutional Ethics Committee at St. John’s Medical College Hospital, Bangalore on 24^th^ July 2021 (Study no. 233/2021).

### Cytokine analysis

Serum samples were thawed and cytokine/chemokine levels were assayed on a multiplex bead array platform. The LegendPlex Human Anti-Virus Response Panel (13-plex) (740390, BioLegend) was utilised according to manufacturer’s instructions with limited modifications as described previously^4^. Data were analyzed using the LegendPlex data analysis software v.8 for Windows.

### IP10 assay [Clinical laboratory test]

IP10 (CXCL10) in sera was measured using the Quantikine ELISA kit (R&D Systems) according to the manufacturer’s instructions, and adopted for use on the automated DS2 ELISA System (Dynex Technologies). Briefly, this is a quantitative sandwich ELISA assay. Microtitre 96-well plates were pre-coated with a monoclonal antibody specific to IP10. With washes between steps, sera and reference material were added, followed by an enzyme-conjugated polyclonal anti-IP10 antibody, then a colorimetric substrate. The concentration of IP10 was estimated by interpolation from the readings of a standard reference curve. The assay was internally verified according to a standard operating procedure for evaluating new assays against pre-set acceptance criteria, including variability, linearity, sensitivity, stability and limits of detection. Continuous performance monitoring was undertaken by plotting the results of Internal Quality Control samples on Levey-Jennings charts and applying the Westgard rules.

### Whole-genome sequencing of SARS-CoV-2

Whole genome sequencing utilised residual SARS-CoV-2 RNA extracted from nose and throat swabs and was performed on GridION (Oxford Nanopore Technology) using version 3 of the ARTIC protocol and bioinformatics pipeline, with lineage calling by pangolin v2. A proportion of total samples was submitted for sequencing based on availability of residual sample.

### Outcomes and predictors

The primary outcome for deterioration was defined as having one of the following: ICU admission or death (ward or ICU setting).

We included four chemokines / cytokines [interferon **γ**-induced protein 10 kDa (IP10), Interleukin 10 (IL-10), Interleukin 6 (IL-6), and Interleukin 8 (IL-8)] and eight blood parameters [white blood cell count, neutrophils, lymphocytes, platelets, creatinine, albumin, bilirubin, and C-reactive protein (CRP)] as potential predictors for the model predicting whether or not patients would deteriorate. For each chemokine / cytokine assayed, data were truncated at 99.5% to avoid the influence of extreme outliers. Patients with any missing predictor data were excluded from the analysis (n=29, 10%). Age was included as the principal demographic variable owing to its previous association with deterioration^12,13^.

### Model development

We used reluctant generalised additive modelling (RGAM) for variable selection and model development in the discovery cohort^14^. Generalised additive models (GAMs) are models that are frequently used to model potential non-linear relationships between outcome and continuous predictors, thereby improving model performance when true relationships between the (transformed) response and a predictor may not be linear^15^. However, GAMs do not work well if the number of potential predictors is large compared to the number of observations. Various sparse additive models, i.e., methods that reduce the number of predictors in the final model, have been proposed, of which RGAMs can be used for binary outcome data^14^. The method has been shown to work well on simulated and real data with small numbers of observations and a large number of features, outperforming other sparse methods such as the lasso and generalised additive model selection with datasets of 100 observations and 200 predictors^14^.

RGAM adapts a reluctant non-linear selection principle, where there is a preference for a linear term over a non-linear term if all else is equal^14^. To implement this principle, RGAM mimics the three-step process of reluctant interaction modelling^16^. First, lasso regression was fitted using the best hyperparameter **λ** for regularization penalty chosen by 10-fold cross-validation. Then, for each predictor, a non-linear feature was constructed using smoothing splines with 4 degrees of freedom and rescaled with gamma=0.6 (default). The rescaling of the non-linear features is undertaken to ensure that non-linearity must be pronounced enough in order to not get penalised out in the final third step.The final step was to refit the model using lasso regression again for variable selection from all the linear and non-linear features generated in the previous step^14^.

### Model validation

The developed predictive models were validated using the validation cohort. The same truncation method was applied to the cytokine variables, and patients with missing data were excluded from the validation cohort (n=32, 18%). We examined the models’ discrimination, calibration, and clinical utility using Decision curve analysis.

Model discrimination, which measures the model’s capability to distinguish patients with the outcome from patients without the outcome, was assessed by the area under the receiver operating characteristic curve (AUROC). It plots sensitivity against 1-specificity for consecutive cut-offs, and normally ranges from 0.5 (no discrimination) to 1 (perfect discrimination).

Model calibration, which measures the agreement between observed data and predictions, was assessed by calibration slope, calibration-in-the-large, and calibration plots^17^. Calibration slope is the regression slope of the linear predictor, and a value close to 1 indicates good calibration. Calibration-in-the-large, which compares the mean predicted risk with the mean observed risk, is the regression intercept by fitting the linear predictor as an offset term: a value close to 0 represents good calibration. Calibration plots show the model predictions against outcome observations, and a line closest to the diagonal reference line indicates good calibration.

Considering that the outcome incidence in the validation cohort was lower than that in the discovery cohort, recalibration was performed by first updating the intercept using calibration-in-the-large. Then, the model was updated by also adjusting the calibration slope^18^. We did not refit the model because, with a relatively small sample size, parsimonious updating methods should be preferred^19^. We found that by updating the intercept only, the models were able to capture the differences in outcome incidence and achieved good calibration scores.

We also performed decision curve analysis, which examines the ‘net benefit’ of prediction models for assessing their clinical utility^20^. The net benefit is calculated as the balance between the likelihood of benefit (correctly identifying true positives) and the likelihood of harm (incorrectly identifying false positives) across a range of threshold probabilities. Decision curve analysis was used in the validation data for all six models, and their net benefit was compared under selected clinically reasonable thresholds. The selection of thresholds was assisted by visualizing model prediction performance (sensitivity, specificity, positive predictive values, negative predictive values) across a range of thresholds (0-0.4) in the validation cohort. There is a direct relationship between the threshold probability and preference of the clinician interpreting or using the decision curve. For example, when using a threshold probability of 0.2, a clinician would be arguing that missing a patient who will deteriorate is 4 times (odds of 1:4) worse than unnecessarily escalating treatment and admitting a patient to ICU. In the case of decisions regarding whether to admit a patient to ICU or not, the threshold probability may vary over time, depending on bed occupancy. Here we only show threshold probabilities up to 0.4, as while acceptable threshold probabilities may vary, it is unlikely that there will be a situation where clinicians would find that missing a patient who will deteriorate is less than 1.5 times worse (odds of 1:1.5) than unnecessarily escalating a treatment. Following classical decision theory, the strategy with the highest expected net benefit should be chosen, irrespective of the size or statistical significance of the benefit^20^.

All analyses were performed using relgam, pROC, rmda, and runway libraries in R (version 3.6).

### Flow cytometry and acquisition

PBMC samples were thawed in a 37°C water bath and washed with 10% heat-inactivated foetal bovine serum in RPMI 1640 medium with GlutaMAX supplement (Life Technologies). Cells were then washed in PBS and stained for viability using live/dead blue (Life Technologies) for 15 minutes at room temperature. Live/dead stained cells were washed and resuspended in FACS buffer prior to surface marker staining. Antibody mastermixes, detailed in Supplementary Table 6 (including the use of Brilliant stain buffer plus (BD) and Trustain Fc receptor blocker (BioLegend) were added to live/dead stained samples and stained for 45 minutes at room temperature. Stained samples were fixed using BD lysing buffer (BD), followed by FACS buffer wash before acquisition on ID7000 (SONY Biotechnology) spectral cytometer. The same cytometer, with the same configuration, was used for the measurements of all the samples. AlignCheck particles (SONY) and 8 peak beads (SONY) were run daily to perform instrument QC and to standardise the cytometer using median fluorescence intensity (MFI) target values, respectively. Flow data (FCS files) were analysed manually using FlowJo software version 10.8 (LLC).

### Flow cytometry data processing

Samples in which all data were absent due to processing errors were excluded from the analysis. To augment the sensitivity and improve the recovery of immunological signals, we eliminated cell populations with low statistical confidence from the analysis. Sample origin was used to partition the dataset and identify differential population recovery between cohorts (India and UK). Our initial step involved a systematic examination of cell counts per population, segmented by the source of the sample.

It is recognized that statistical power and confidence diminish as cell counts approach zero. Low cell counts may introduce sampling noise, sources of which may include batch effects and differences in sample preparation. Such noise tends to dominate the signal at minimal cell counts. Consequently, populations exhibiting low counts were systematically identified as candidates for removal based on their average counts across all samples within each cohort. A population was retained if its average cell count surpassed a predefined threshold of confidence. Conversely, if a cell population demonstrated consistently low counts across all cohorts, it was excluded from further analysis. This decision was based on the rationale that such populations do not contribute meaningful variation and may introduce more noise than signal into subsequent analyses.

This selection process led to the inclusion of 1489 and 1455 populations from the India and UK cohorts, respectively, a total of 1556 retained populations out of the possible 1924 measured across the entire panel. This approach ensures that only cell populations contributing robust and statistically significant signals are included in the analysis, enhancing overall quality and reliability.

### Statistical analysis for immune-profiling

All analyses of immune-profiling data were performed in R version 4.3.1. Spearman’s R correlations of immune populations with IP-10 and Kendall’s tau correlations of immune populations with ordinal disease severity were performed with rstatix v 0.7.2 and Kendall v 2.2.1 packages, respectively. P-values were corrected with the Benjamini-Hochberg method. For non-redundant analysis of correlating immune lineages, the mean p-value and test statistic were reported for any equivalent lineages measured in more than one antibody panel. This was likewise applied to cell type-phenotype relationships depicted in chord diagrams, which were additionally condensed (calculating median p-values and test statistics) to summarise trends in single surface markers and to avoid double counting of inverse immune phenotypes. Chord diagrams were generated using circlize v 0.4.16^21^. For summary of populations correlating with both IP-10 and disease severity, arbitrary correlation units were calculated as the mean of the test statistic from co-correlating Kendall’s tau and Spearman’s R associations.

## RESULTS

### Patient cohorts and characteristics

This study presents data based on serum cytokine and chemokine measurements in three distinct cohorts analysed sequentially between 2020 and 2022, that, together with controls comprised 659 persons. The three cohorts comprised: i) a first-wave SARS-CoV-2-naive, pre-vaccine UK cohort; ii) pauci-vaccinated patients infected with the B.1.1.7 variant in the UK; and iii) a 2022 India part-vaccinated cohort (1-3 doses) who received immunomodulators as standard, contingent on oxygen threshold, but among which a proportion nevertheless deteriorated.

In the UK, serum was collected from 454 patients, PCR-confirmed to have COVID-19 at hospital admission. Individuals who entered the study between March to November 2020 were assigned to the Discovery cohort [n=278] and those sampled between January to March 2021 formed the Validation cohort [n=176] (Fig. 1a). Demographics including age, gender, body mass index and ethnicities (non-Caucasian: Discovery 51.6%; Validation 49.4%) were similar for both groups, as was the prevalence of malignancy (Discovery 8.3%; Validation 9.7%), which is independently associated with mortality, irrespective of age (Supplementary Table 1)^22^. Nonetheless, the cohorts could not be perfectly matched with the representation of Black individuals and diabetes being significantly greater in the Discovery cohort, whereas Latin individuals (albeit a very small percentage) and ischaemic heart disease were over-represented in the Validation cohort (Supplementary Table 1). A proportion of virus isolates were submitted for sequencing: whereas no dominant variant was evident for the Discovery cohort, 99% of sequences in the Validation cohort were of the alpha (formerly B.1.1.7) variant, consistent with the epidemiological shift across the UK (Fig. 1b).

**Figure 1.**
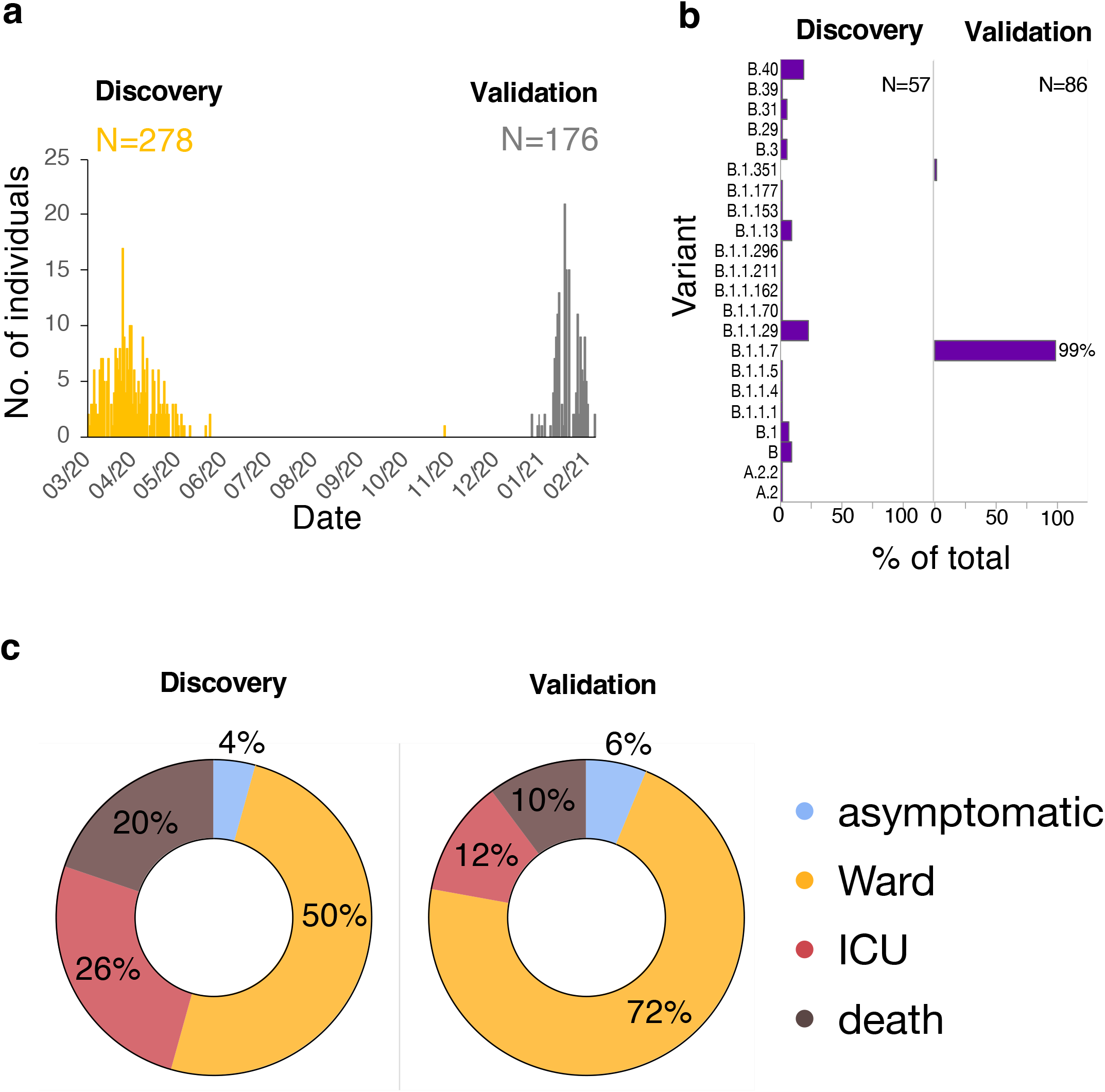
Clinical characteristics of Discovery and Validation cohorts. a, Patients confirmed to have SARS-CoV-2 were recruited between March to November 2020 [Discovery Cohort; n=278] and January to March 2021 [Validation cohort; n=176]. b, A proportion of SARS-CoV-2 isolates were sequenced; histograms demonstrate the percentage of each variant in the Discovery and Validation cohorts. c, 30-day outcome data is shown for each cohort; no data were missing. Patients admitted to the ward were differentiated into those who were ‘asymptomatic’ (identified on routine screening) or symptomatic and admitted to an inpatient non-ICU bed.

Patients were subdivided into the following groups based on their 30-day outcome; asymptomatic (incidental diagnosis on routine screening), admission to the ward, admission to intensive care (ICU) or death (Fig. 1c). All patients had outcome data recorded. Fewer individuals deteriorated (composite of ICU admission and or death) in the Validation cohort (22%) versus the Discovery cohort (46%), reflecting improvements of care including re-purposed therapeutics, most notably steroids. Deterioration was not related to time from symptom onset to admission blood sample (Supplementary Fig. 1).

### Parameters anticipating clinical deterioration

Sixteen clinical laboratory and cytokine/chemokine measurements were assessed as candidate predictors of downstream clinical symptoms. They were selected based on extensive literature review and our own published studies^4–8,23,24^. Given their prior negative association with in-hospital outcomes, we additionally evaluated age, BMI and admission National Early Warning Score (NEWS2)^12,13,25–27^ (Fig. 2a; Supplementary Table 2). All clinical variables with missingness greater than 10% (bilirubin, glucose, alanine aminotransferase, ferritin, urea, D-dimer, Troponin) were excluded, as were cytokines/chemokines (IFN-γ, IFNλ1, IFNλ2/3) for which the dynamic ranges of available assays were mostly not suitable *vis-à-vis* typical serum concentrations for COVID-19 patients or healthy controls (Supplementary Table 2).

**Figure 2.**
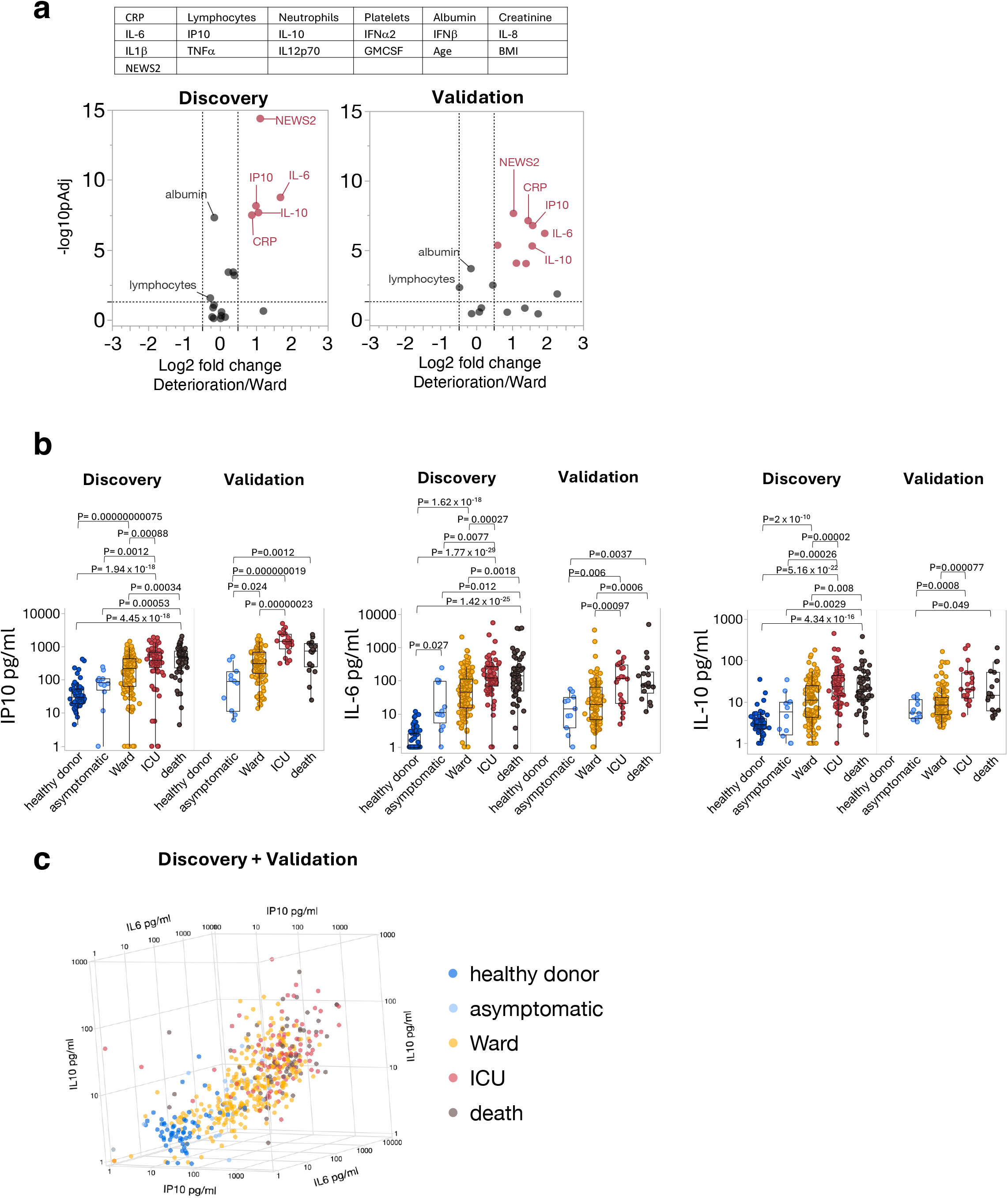
A triad of IP10, IL-6 and IL-10, measured at admission, discriminate patients who deteriorate. a, Volcano plots show 19 variables (age, BMI, NEWS2 score, clinical laboratory analytes with <10% missing values and cytokine parameters), measured at the point of admission, comparing patients who received ward level care versus those who deteriorated [intensive care admission and or death] in the Discovery (left volcano; ward n=151; deterioration n=127) and Validation (right volcano; ward n=137; deterioration n=39) cohorts. A prediction horizon for deterioration was set at 30 days. Statistical analysis was performed using the two-tailed Mann–Whitney test. Statistically significant parameters are marked in red (Log2fold change >0.5 or <-0.5 and Benjamini–Hochberg (BH) corrected p-value < 0.05). b, IP10, IL-6 and IL-10 levels are compared between the groups for each cohort. P-values were generated by a Kruskal–Wallis test with Dunn’s post hoc correction. Box plots show median, first quartile (Q1) and 3^rd^ quartile (Q3) and whiskers extend between Q1-1.5 x IQR (interquartile range) and Q3+1.5 x IQR. Values below the assay limit of quantification were set as 1 pg/ml. c, Relationship between IP10, IL-6 and IL-10 in all patients and healthy donors (Discovery and Validation cohorts combined). Patients are grouped by the most severe outcome within 30 days [Healthy donor n=61, Asymptomatic n=23, Ward n=265, ICU n=93, Death n=73].

Fig. 2a displays a volcano plot based on how each parameter, measured either at between 0 hours and 24h post-admission (94%) or within 48h of admission (6%) segregated with ward-level care *versus* deterioration (intensive care and/or death) over a 30-day window in the Discovery cohort (left volcano plot; ward n=151; deterioration n=127) and Validation cohort (right volcano plot; ward n=137; deterioration n=39), respectively. Significant associations with severity were shown in both cohorts for the NEWS2 score^28^, which is a validated tool used to detect clinical deterioration; for CRP; and for IP10, IL-6, and IL-10. Indeed, individual donor serum measurements (Fig. 2b,c) showed consistent, significantly greater levels of each of the three analytes in those persons progressing to ICU *versus* those requiring only ward-care. Moreover, within the IP10, IL-6, and IL-10 triad, the most significant discriminator of ward to ICU was IP10 (Fig. 2b), seemingly consistent with several other, independent reports published following our initial COVID-IP study that likewise claimed that IP10 levels were predictive of subsequent deterioration in COVID-19^29–33^.

By contrast, among the clinical laboratory measures, validated prognostic associations with subsequent deterioration were evident only for CRP and NEWS2, consistent with the volcano plot (Fig. 2a), and for neutrophils for which the associations were weaker (Supplementary Fig. 2). Likewise, there was no consistent and significant association with measurements of IL-12p70, IL-1β, TNFα, GMCSF, and Type I interferons, although IL-8 levels were significantly higher in death versus ward patients in both Discovery and Validation cohorts (Supplementary Fig. 2).

### Development and validation of a predictive model of COVID-19 deterioration in the UK

We next developed multivariable predictive models for deterioration to formally assess whether addition of easily measurable immune analytes to standard laboratory tests might increase test accuracy to identify patients at risk of deterioration. Sensitive to the pressures in a pandemic setting, we sought test parameters that can be measured within an NHS laboratory, or in equivalent clinical laboratories in other countries, negating any requirement for a clinician’s time and input to calculate the score. Specifically, we excluded parameters, e.g., NEWS2, that include physiological measurements which are commonly recorded manually on paper or on e-noting systems, neither of which routinely communicates with laboratory software. Age was included as the principal demographic variable owing to its previous association with deterioration^12,13,27^.

Reluctant generalised additive modelling (RGAM) was used for variable selection and model development in the Discovery cohort (see STAR Methods). Based on the literature, current clinical practice, and our own data, a suite of eight routinely measured laboratory blood parameters (white cell count, neutrophils, lymphocytes, platelets, creatinine, albumin, bilirubin, CRP) and four chemokines /cytokines (IP10, IL-10, IL-6, and IL-8) were selected as predictors. Note that for each chemokine or cytokine, data were truncated at 99.5% to avoid influence of extreme outliers. Patients with any missing predictor data were excluded from the analysis (n=29, 10%).

Six different models predicting deterioration were built: 1) included chemokine and cytokine predictors only; 2) included the clinical laboratory blood predictors only; 3) included all clinical laboratory blood and chemokine/cytokine predictors; 4) included clinical laboratory blood predictors and IP10; 5) included clinical laboratory blood predictors plus IL-6; and 6) included IP10 alone. Note, we compared the utility of incorporating IL-6 with the clinical laboratory blood predictors because of the implications of heightened IL-6 levels in contributing to COVID-19 severity^4,6^, and their efficacious amelioration in some cases by clinical use of anti-IL-6 antibody, tocilizumab^34,35^. We chose to evaluate IP10 alone based on its compelling association with deterioration (see above) and given that at the time of sampling, a neighbouring pathology laboratory had verified the IP10 ELISA to ISO 15189 standards (a test which has since become UKAS accredited), to aid the management of complex patients with ongoing healthcare needs owing to COVID-19.

Discrimination and calibration metrics for six models are shown in Table 1. The data establish that AUROC values were comparable for all chemokine/cytokine predictors or IP10 in isolation, compared to routinely employed clinical laboratory parameters, whilst the combination of all cytokines/chemokines or solely IP10 with clinical laboratory parameters achieved the highest AUROC performance (clinical lab and cytokines, 0.88; 95%CI 0.82-0.95; clinical lab and IP10, 0.88; 95% CI 0.82-0.94) (Fig 3a; Table 1). Addition of IL-6 alone to clinical laboratory markers, however, did not improve model performance (0.80; 95% CI 0.72-0.89) compared to clinical laboratory parameters alone (0.79; 95% CI 0.71-0.88) (Fig. 3a; Table 1).

**Figure 3.**
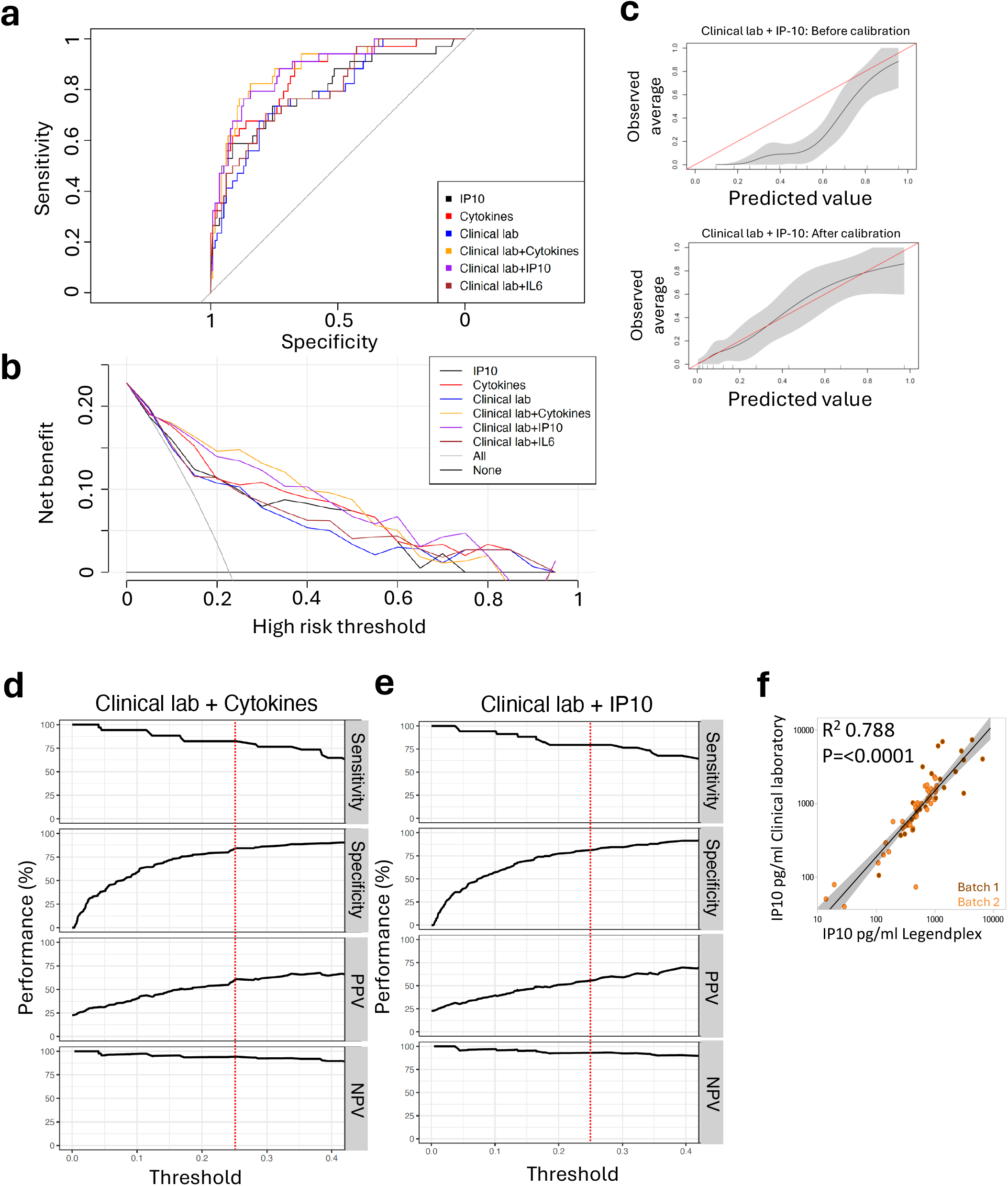
IP10 improves prognostication of deterioration. a, Comparison of area under the receiver operating characteristic curve (AUROC) among six models in the validation cohort. Models are recalibrated to the validation data. b, Decision curve analysis across eight models in the validation cohort. Net benefit is plotted for each model compared with the treat-all and treat-none approaches. Models are recalibrated to the validation data. c, Calibration plot for cytokine and IP10 model, before (left) and after (right) recalibration in the validation cohort. Calibration curve is shown using locally weighted smoothing (LOESS) function. Gray shade shows the 2 standard error (RE) region for the curve. Red line is the diagonal reference line. Sensitivity, specificity, positive predictive values (PPVs) and negative predictive values (NPVs) of the d, clinical lab and cytokines or e, clinical lab and IP10 model in the validation cohort, according to the full range of probability thresholds for the model prediction. Red dotted lines show a threshold of 0.25. f, IP10 was determined from 64 admission samples (in two separate batches, differentiated by colour) on a UKAS accredited IP10 ELISA in routine use within a pathology laboratory versus a commercially available multiplex bead array carried out in our laboratory, as per manufacturer’s instructions. A linear regression line with 95% confidence intervals [shaded area] is shown. R^2^ and P values are shown on the graph.

**Table 1.** Discrimination performance in the UK Discovery cohort and discrimination and calibration performance in the UK Validation cohort.

| Discovery |  | Validation |  |  |
| --- | --- | --- | --- | --- |
|  | AUC | AUC | Calibration-in-the-large | Calibration slope |
| IP10 | 0.68(0.62, 0.75) | 0.80(0.70, 0.89) | -1.90(-2.50, -1.38) | 1.14(0.74, 1.58) |
| Cytokines | 0.74(0.68, 0.80) | 0.85(0.78, 0.92) | -1.65(-2.23, -1.16) | 1.73(1.18, 2.38) |
| Clinical lab | 0.79(0.73, 0.84) | 0.79(0.71, 0.88) | -1.04(-1.49, -0.62) | 1.69(1.05, 2.45) |
| Clinical lab + Cytokines | 0.81(0.76, 0.86) | 0.88(0.82, 0.95) | -1.74(-2.39, -1.20) | 1.88(1.28, 2.61) |
| Clinical lab + IL-6 | 0.79(0.74, 0.85) | 0.80(0.72, 0.89) | -0.97(-1.43, -0.54) | 1.86(1.18, 2.67) |
| Clinical lab + IP10 | 0.81(0.75, 0.86) | 0.88(0.82, 0.94) | -1.82(-2.48, -1.26) | 1.81(1.23, 2.53) |

In considering whether there might be true benefit to patients, the decision curve analysis in the validation cohort showed that all models had higher net benefit than the treat-all or treat-no one strategies across a range of threshold probabilities, but that none of the models had consistently higher net benefit than other models (Fig. 3b). Given that the calibration metrics for the models were not ideal (e.g., for the clinical lab+IP10 model, calibration-in-the-large was −1.82 (95%CI −2.48-1.26) and the calibration slope was 1.81 (95% CI 1.23-2.53) (Table 1), all models were recalibrated to the validation data before generating the decision curves (Fig. 3a-b). Recalibration generally led to similar predicted and observed values as shown for the clinical lab + IP10 model (Fig. 3c). For the same model with clinical lab and IP-10 measurements included, recalibration led to a net reclassification improvement of 56% (Table 2). More specifically, recalibration model 4 gave 101 out of 115 (88%) patients who did not deteriorate a lower predicted risk of deterioration.

**Table 2.** Recalibration table for clinical lab and IP10 model in the validation cohort.

| Prediction model | Recalibrated model |  |  | Reclassified |  | Net correctly reclassified (%) |
| --- | --- | --- | --- | --- | --- | --- |
|  | <10% | 10-35% | >35% | Move up | Move down |  |
| Non-deterioration (N=115) |  |  |  |  |  |  |
| <10% | 1 | 0 | 0 | 0 | 101 | 88% |
| 10-35% | 45 | 0 | 0 |  |  |  |
| >35% | 20 | 36 | 13 |  |  |  |
| Deterioration (N=34) |  |  |  |  |  |  |
| <10% | 0 | 0 | 0 | 0 | 11 | -32% |
| 10-35% | 2 | 0 | 0 |  |  |  |
| >35% | 0 | 9 | 23 |  |  |  |
| Net reclassification improvement |  |  |  |  |  | 56% |

The sensitivity, specificity, PPV, and NPV of the clinical laboratory tests augmented by all cytokines/chemokines or augmented solely by IP10 across the full range of evaluated probability thresholds (0-0.4) for the model prediction in the validation cohort is shown (Fig, 3d-e). A threshold of 0.25 was chosen since this reflects an acceptable clinical setting *de facto* in which sensitivity (the capacity to predict deterioration) is valued over specificity, i.e., those falsely considered to be at-risk for deterioration, and hence treated as such, are less important than potentially missing an at-risk patient. At this threshold (demarcated by the red dashed line), sensitivity and specificity each reached 80% for both models, a relatively high performance level (Fig. 3d,e).

### Short-term feasibility of model utility in clinical practice

Given the retrospective capacity of deterioration to be predicted by a combination of IP10 and routine clinical laboratory measurements, we assessed the practical capacity to apply it in the clinic in the immediate term. Thus, 64 serum samples were subject to a clinically validated, UKAS accredited, IP10 assay routinely run over two batches within a UK pathology laboratory. IP10 levels were used in clinical practice on-site to guide management of complex COVID-19 cases. Excellent concordance was seen between the clinical assay and the research assay that we had used to measure IP10, with negligible batch effect (R^2^=0.788, P<0.0001) (Fig. 3f). This thereby established a clinical pathway by which to implement the model with immediate effect.

We next considered it important to investigate model performance and the significance of at-admission IP10 levels in more recent clinical practice, in which COVID-19 cohorts are more heterogeneous than those earlier in the pandemic by many criteria including variable amounts of prescribed vaccination, variable amounts of vaccination *via* natural exposure to virus, diversity of prevailing virus variants, and widespread adoption of immunomodulatory therapeutics, including dexamethasone. To this end, we examined a cohort of 88 persons from Bangalore, India who were hospitalised between January 2022-February 2023 either because they were diagnosed with COVID-19 or because they were already hospitalised for another condition but tested positive for SARS-CoV-2. In addition, 100 patients without COVID-19 were recruited from the same hospital (Supplementary Table 3).

Given that admission blood tests in the India cohort were not performed routinely, we were unable to assess models that incorporated clinical laboratory parameters, but we could assess cytokines/chemokines or IP10 alone, as shown (Table 3; Fig 4a-c). AUROC values were substantially worse in this validation cohort than the validation cohort from the UK for both the IP10 only model (India: 0.57, 95%CI 0.43-0.71; UK: 0.80, 95%CI 0.70-0.89) and the cytokines only model (India: 0.57, 95%CI 0.43-0.71; UK 0.85, 95%CI 0.78-0.92) (Tables 1, 3). The decision curve also indicated that the net benefits of using these models were much less than in the UK cohort (Fig. 3b, Fig. 4a). However, against the outcome of death alone, models based on cytokines or IP10 showed improved performance, compared to UK Discovery and Validation cohorts (Supplementary Table 4, Supplementary Figs. 3, 4). Of the nine deaths, eight had severely deteriorated on ICU, and they showed significantly higher levels of IP10, IL-6 and IL-8, with a trend for IL-10, when compared to patients in the non-COVID-19 group (Fig. 4d,e).

**Figure 4.**
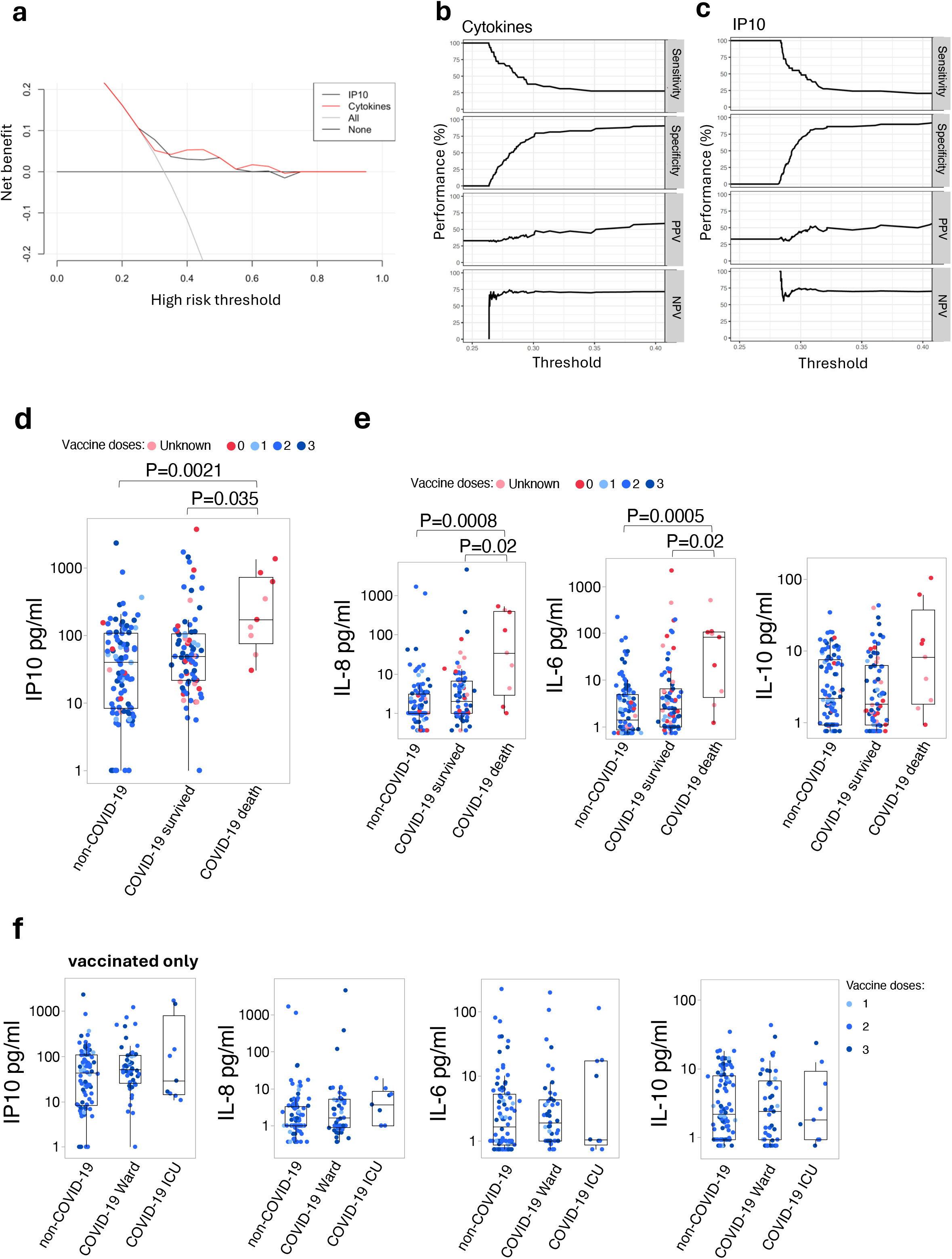
IP10 remains a signature of death within a tertiary hospital in India. a, Decision curve analysis across four models in the India Validation cohort. Net benefit is plotted for each model compared with the treat-all and treat-none approaches. Models are recalibrated to the validation data. Sensitivity, specificity, positive predictive values (PPVs) and negative predictive values (NPVs) of b, cytokines or c, IP10 models in the India Validation cohort, according to the full range of probability thresholds for the model prediction. Serum d, IP10 and e, IL-8, IL-6 and IL-10 levels in admission samples from patients at a tertiary hospital in India. Spot colours correspond to number of prior vaccine doses received. P-values were generated by a Kruskal–Wallis test with Dunn’s post hoc correction. Box plots show median, first quartile (Q1) and 3^rd^ quartile (Q3) and whiskers extend between Q1-1.5 x IQR (interquartile range) and Q3+1.5 x IQR. f, as in panel e but for patients with evidence of prior vaccination.

**Table 3.** Discrimination and calibration performance in the India validation cohort.

|  | Discovery (UK) | Validation (India) |  |  |
| --- | --- | --- | --- | --- |
|  | AUC | AUC | Calibration-in-the-large | Calibration slope |
| IP10 | 0.68(0.62, 0.75) | 0.57(0.43, 0.71) | -0.47(-1.00, 0.07) | 0.52(-0.04, 1.16) |
| Cytokines | 0.74(0.68, 0.80) | 0.57(0.43, 0.71) | -0.07(-0.80, 0.74) | 0.79(0.08, 1.58) |

Of note, all nine COVID-19 non-survivors, had either no evidence of prior vaccination or evidence that they were not vaccinated (Fig. 4d,e). By contrast, most of the patients surviving COVID-19 had been vaccinated (Fig. 4d,e), and in the timeframe of admission or shortly thereafter, the levels of IP10, IL-6, IL-8, and IL-10 in vaccinated individuals showed no capacity to discriminate ICU patients from either COVID-19 ward patients or patients without SARS-CoV-2, many of whom were younger than the COVID-19 patients (Fig. 4d,f; Supplementary Table 3). Of note, the IP10 levels were comparable to those in the normal range of healthy control donors and asymptomatic patients displayed in Fig. 2b (above). In sum, our data support the continued utility of immune biomarkers at a later stage in the pandemic for the prediction of subsequent terminal deterioration, even where and when at-admission blood tests are not routinely available, and suggest that this may be primarily beneficial for the management of unvaccinated patients.

### Immunological correlates of IP10 and COVID-19 severity

Our study to this point has validated the association of high at-admission IP10 levels with subsequent and severe clinical deterioration in three independent and highly diverse settings, to the point that measures of IP10 levels have added demonstrable predictive value when added to routine clinical laboratory tests. This in turn raises the question as to what high IP10 levels might reflect in relation to seeking a better understanding of disease causation.

To this end, we applied high-content, high-throughput spectral flow cytometry-based immune-profiling technologies that are substantially and substantively advanced relative to those used in original profilings of patients with COVID-19^4^, and which discriminate >1800 immune cell parameters, including cell types and activation states of T, B and myeloid lineages (see STAR Methods). We retrieved stored PBMC from 34 of 88 COVID-19-positive patients for whom paired serum samples were available, and 51 SARS-CoV-2-negative patients from the Bangalore cohort. For comparison, we re-examined samples from 17 donors across a range of COVID-19 severities, examined in 2020 as part of the COVID-IP cohort^4^ by high-throughput flow cytometry that was state-of-the-art at that time (Fig. 5a and Supplementary Table 5), thereby establishing consistency of our approaches. First, we determined cellular immune correlates of high IP10 levels in a continuous variation model; second, we determined cellular immune correlates of COVID-19 severity, measured as ordinal outputs; and third, we asked if there was a substantial overlap of the two.

**Figure 5.**
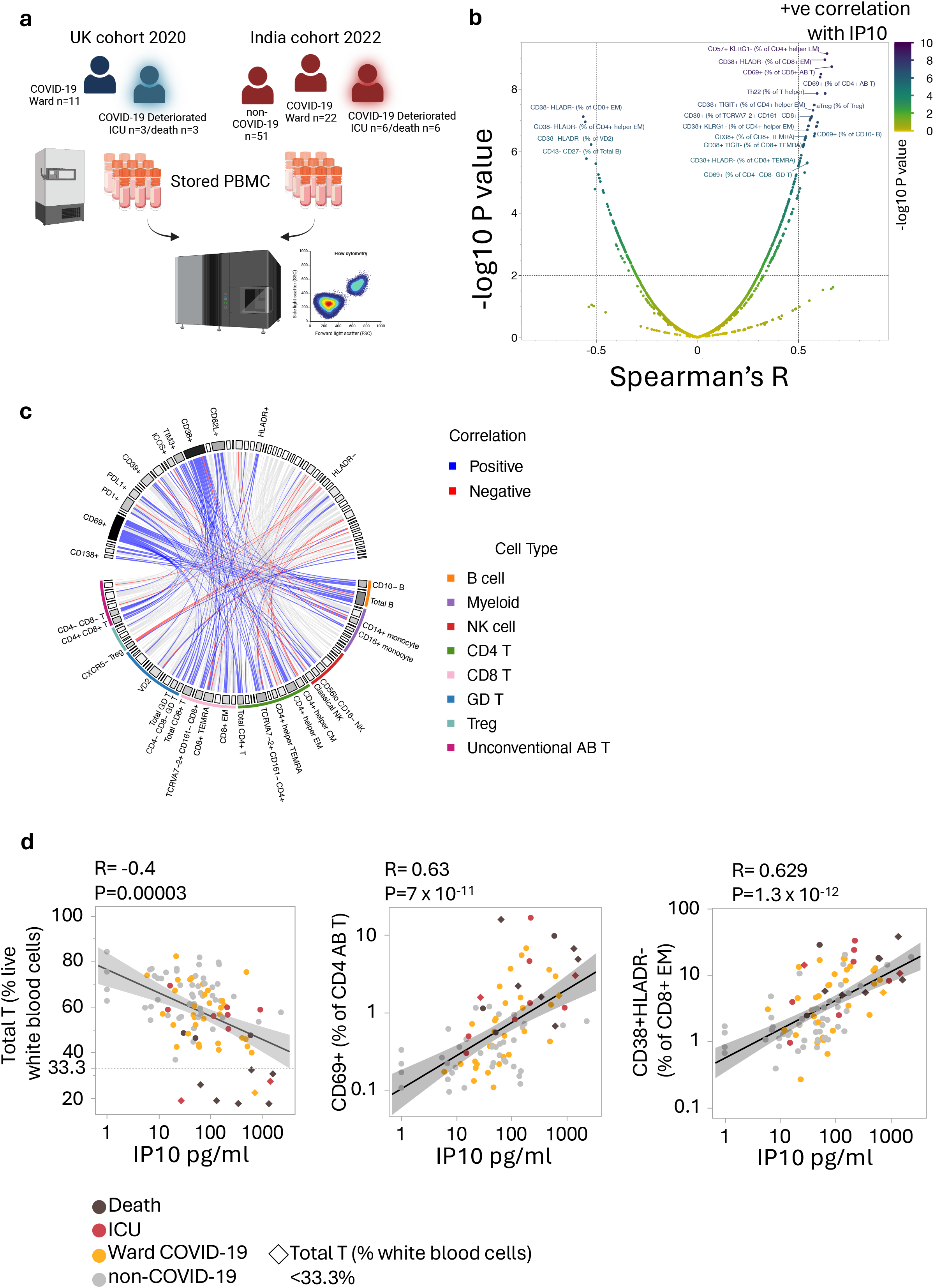
High IP10 encompasses a cellular signature of severe COVID-19. a, Outline of experimental design. Banked PBMC samples were retrieved from two geographically distinct cohorts, each with a high proportion of in-patient deterioration, and cellular traits were assessed by multi-parameter spectral flow cytometry. Created with Biorender.com. b, Volcano plot shows immune cell correlations with serum IP10 in combined samples from India 2022 (COVID-19 + non-COVID-19, n=85) and UK 2020 (COVID-19, n=17) cohorts. Correlations measured by Spearman’s R with p-value correction by the Benjamini-Hochberg method (FDR). c, Non-redundant cell surface phenotype-parent cell relationships positively correlated (p<0.01) with IP10. Links represent Spearman R weighting, link colours represent Spearman R directionality in statistically significant (p<0.01) relationships, and sector colours represent relative enrichment (Spearman weighting + number of associations) of indicated markers and cell types. d, Representative examples of significantly correlated immune populations. Diamond symbols demarcate patient samples in which Total T cell (% live white blood cells) frequencies fell beneath 33% of all live white blood cells. A linear regression line with 95% confidence intervals [shaded area] is shown. Spearman’s R and P values are shown on the graphs.

A volcano plot for the first correlation showed that donors with high IP10 levels were enriched in many T cell states, mostly unified by their expressing the activation markers, CD38 and CD69, whereas donors with lower IP10 levels were enriched in CD38^(-)^ T cells (Fig. 5b-c). Moreover, significant phenotypic differences in T cells (e.g., frequency of CD4 T cells that were CD69^+^ and frequency of CD8 effector memory T cells that were CD38^+^HLA-DR^(-)^) were observed in patients with high IP10 levels, including in cases where the T cell pool was already cytopenic (Fig. 5d). In short, high IP10 correlated at admission or closely thereafter with a T cell compartment that was coincidentally highly activated (CD38^+^, CD69^+^, and, to a lesser extent, PD1^+^) and actively depleted, which can be assumed to be a grossly dysfunctional state. Notwithstanding the preponderance of T cell phenotypic changes, positive correlations were also observed for B cell and NK cell phenotypes (e.g., CD69^+^), as illustrated by the depiction of significant, non-redundant relationships between cell surface markers and parent cell types each significantly correlated wih high IP10 levels (Fig. 5c).

When cellular immune correlates of severity were assessed, a similar pattern was seen, with enrichments for T cell phenotypes spanning, e.g., CD4, CD8, effector-memory T (T_EM_) and γδ cells, as well as NK cells expressing CD69 and CD38, respectively. Additionally, there was over-representation of defined B cell states, particularly PD1^+^ or PD-L1^+^ (Fig. 6a-c), with PD1 expression in many settings collectively contributing to stronger relative enrichment (Spearman weighting + number of associations) than was the case for IP10 associations (Fig. 6a-c). Akin to the association with high IP10 levels, T cytopenia correlated with severity (Fig. 6c), consistent with other reports^8,36,37^.

**Figure 6.**
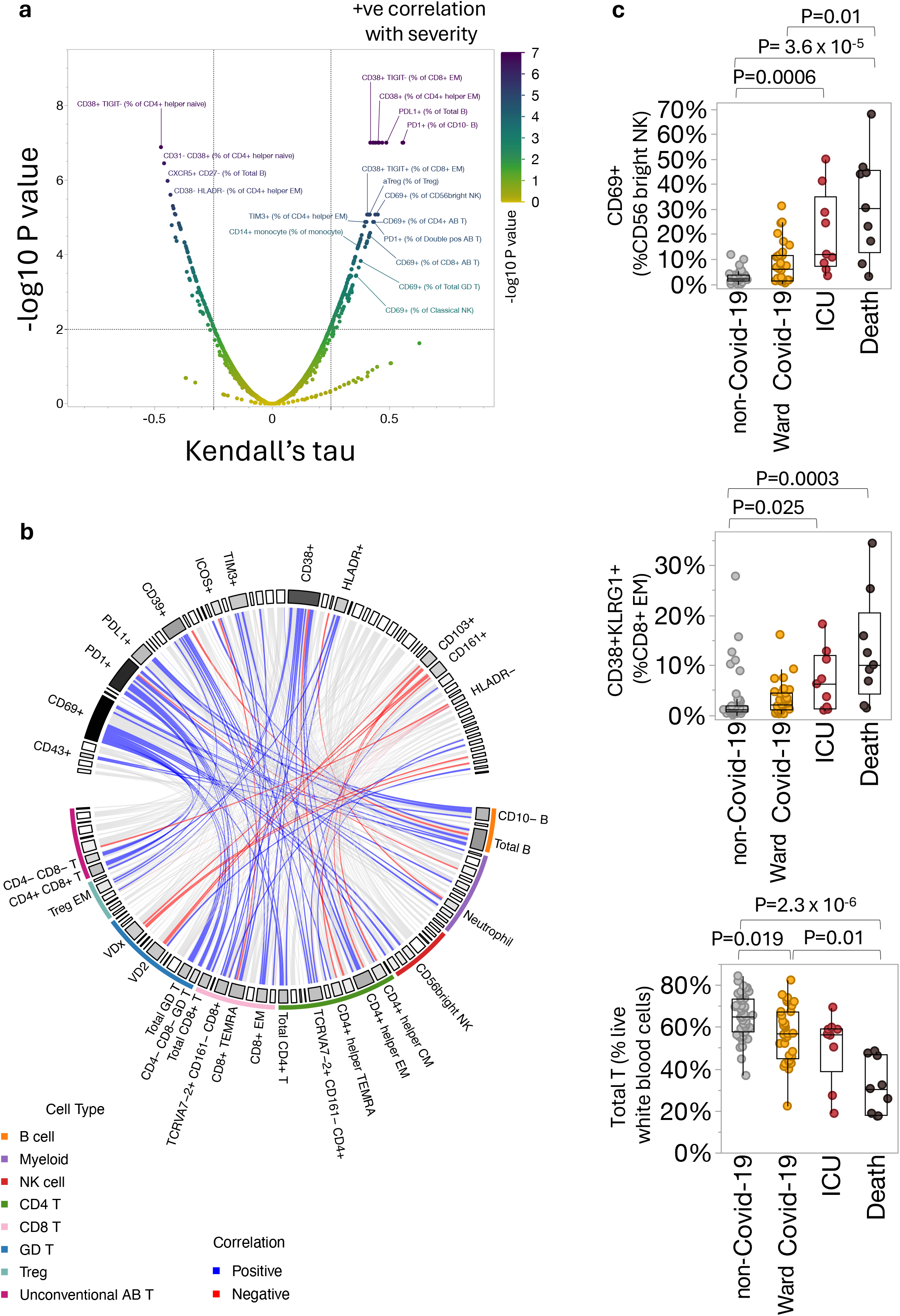
A cellular signature of COVID-19 severity. a, Volcano plot shows immune cell correlations with disease severity (ranked as non-COVID-19, Ward COVID-19, ICU and death) in combined samples from India 2022 (Ward COVID-19 + non-COVID-19, n=85) and UK 2020 (COVID-19, n=17) cohorts. Correlations measured by Kendall’s tau with p-value correction by the Benjamini-Hochberg method (FDR). b, Non-redundant cell surface phenotype-parent cell relationships positively correlated (p<0.01) with disease severity. Links represent Kendall tau weighting, link colours represent Kendall tau directionality in statistically significant (p<0.01) relationships, and sector colours represent relative enrichment (Kendall tau weighting + number of associations) of indicated markers and cell types. c, Representative examples of significantly correlated immune populations in a. P-values were generated by a Kruskal–Wallis test with Dunn’s post hoc correction. Box plots show median, first quartile (Q1) and 3^rd^ quartile (Q3) and whiskers extend between Q1-1.5 x IQR (interquartile range) and Q3+1.5 x IQR.

Examining the overlaps, we found that approximately two-thirds of all statistically significant immunological correlates of severity also correlated significantly with IP10 levels, while the reciprocal was true for over half the correlates with IP10 levels (Fig. 7a), validating the hypothesis that IP10 can be a biologically meaningful prognostic biomarker because of its multiparameter association with subsequent clinical detioration. The dominant cell states among the overlapping correlates were those associated with activation / exhaustion (e.g., CD69, CD38, HLA-DR and TIM3, and T cytopenia), particularly within CD4 helper and CD8 T_EM_ populations. This suggests that among other things, highly dysregulated and actively depleted T cells at hospital admission are both reflected by high IP10 levels and potentially contributory to subsequent clinical deterioration. Additionally, the lineage frequencies most commonly co-associated with IP10 levels and disease severity included monocytes, Tregs, B cell subtypes and CD4 helper types (e.g., Th1, Th2, Th22 [see Fig. 5b, above]). Nonetheless, several PD1-expressing cell subsets were significantly associated with severity but not with IP10 levels (Fig. 7b; yellow data-points), emphasising that high IP10 levels will not capture all correlates of severity, as is most commonly true for other candidate prognostic biomarkers.

**Figure 7.**
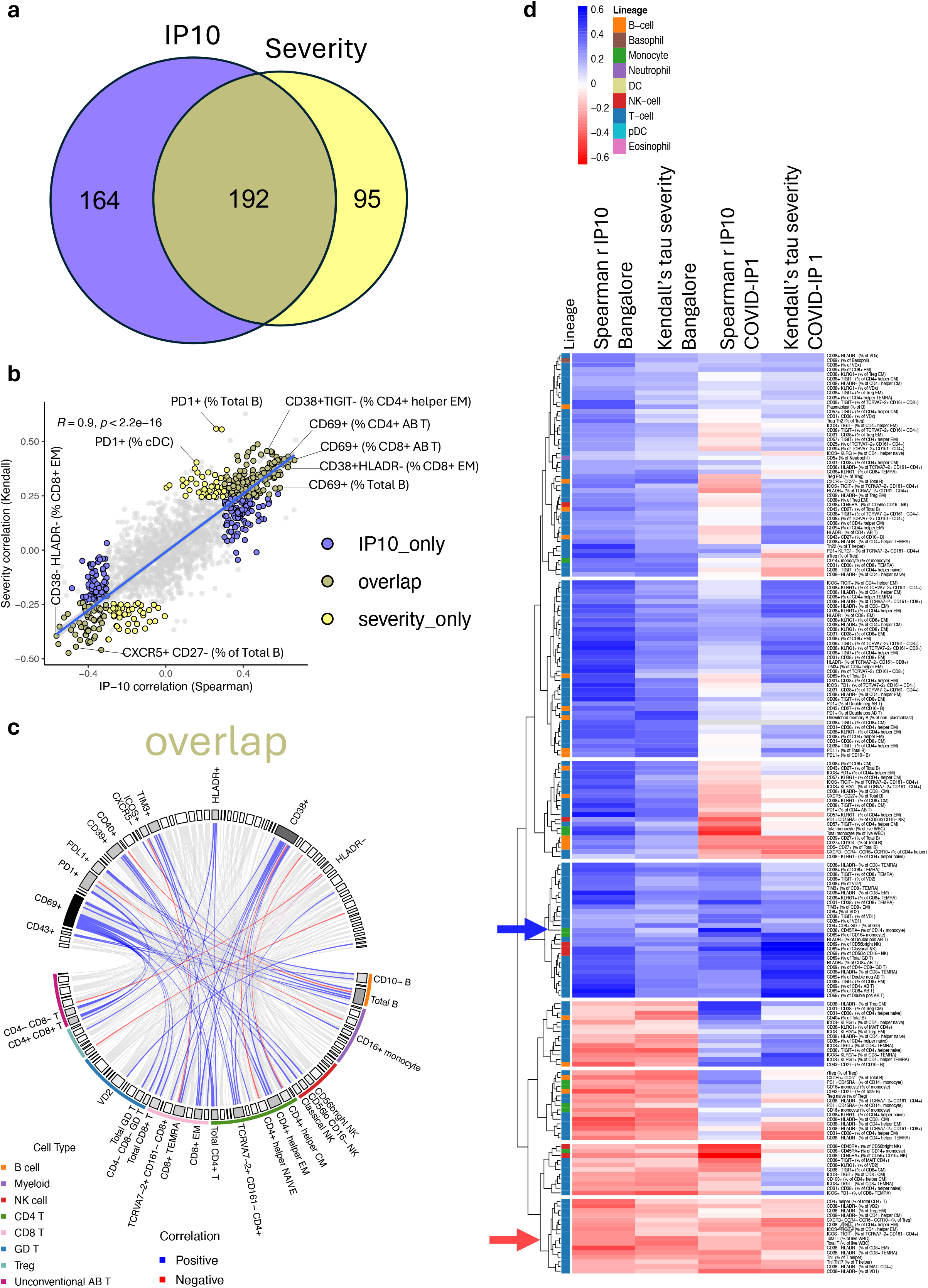
Significant traits correlating with IP10 and severity. a, Overlap in significant (p<0.01) immune cell correlations with IP10 levels (Spearman’s R) and/or disease severity (Kendall’s tau) in India 2022 (Ward COVID-19 + non-COVID-19) and UK 2020 (COVID-19) cohorts. P-value correction by the Benjamini-Hochberg method (FDR). b, Directionality of correlations with both IP10 and severity. c, Non-redundant cell surface phenotype-parent cell relationships positively correlated (p<0.01) with both IP10 and disease severity. Links represent arbitrary correlation weighting, link colours represent correlation directionality in statistically significant (p<0.01) relationships, and sector colours represent relative enrichment (correlation weighting + number of associations) of indicated markers and cell types. d, Immune cell correlations with IP10 levels (Spearman’s R) and/or disease severity (Kendall’s tau) in India 2022 (Ward COVID-19 + non-COVID-19) and UK 2020 (COVID-19) cohorts. All populations were significantly correlated with both IP10 and severity in the combined cohort (p-value<0.01 after correction by the Benjamini-Hochberg method (FDR)). Blue arrow shows a set of parameters which positively associated with severity and IP10 in India and London cohorts. Red arrow shows a set of parameters which negatively associated with severity and IP10 in India and London cohorts.

### Immune correlates across cohorts

To understand how these co-correlates of IP10 and severity persisted between geographically and temporally distinct cohorts, we compared their correlations with the two measures when the India and London cohorts were segregated. Unsupervised clustering identified sets of these co-correlates by their shared or distinct association with the two cohorts (Fig. 7d; significant positive correlations are shown in blue and negative corelations in red). Importantly, despite the considerable differences that the two cohorts presented, many co-correlates displayed similar patterns or association across them (Fig. 7d; note, the red-arrowed group of parameters compose negative correlates across severity and IP10 and geographical setting, while the blue-arrowed group composes positive correlates). Thus, just as at-admission IP10 levels retained their capacity to predict the most severe sequalae of infection, their context remained largely consistent, as reflected by the correlates in common.

Nonetheless, there were exceptions (Fig. 7d). For example, a set of markers including CD38^+^ naïve helper CD4 T cells and CD40^+^ B cells showed marked positive correlation with disease severity and IP10 in the London cohort, but negative correlation in the India cohort (Fig. 7d). The opposite was true of a set of parameters which positively associated with severity and IP10 in the India cohort but showed mild to no association in London patients, including total monocyte frequencies and CD38^+^ CD8^+^ CM cells (Fig. 7d). These differential associations possibly reflect influences on the immune system of geographic effects, including differential exposure to prescribed steroids, and/or pre-exposure legacies including infection and vaccination. By filtering out such differential associations, one can identify the core associations of subsequent severity that are likewise associated with high IP10 levels: from the data presented, those are overwhelmingly composed by T cell phenotypes (Fig. 7d).

## DISCUSSION

We have shown that IP10 measurements combined with routinely employed clinical laboratory blood tests at or very close to the point of hospital admission offer a simple and improved means to prognosticate subsequent clinical deterioration from COVID-19 over a 30-day window in independent Discovery and Validation cohorts in the U.K. At a threshold of 0.25, IP10-plus-laboratory tests showed values of 0.8 for sensivity and specificity. Our findings are supported by other reports appearing after our COVID-IP study that independently attested to the value of IP10 measurements in predicting clinical deterioration in COVID-19^29–33^. In this regard, IP10 measurements seemingly offer insights into the pathology of COVID-19 that are less likely to be offered by physiologic measurements that are often generic across very many disease settings. Besides these specific benefits to clinical management of COVID-19, our findings underline the potential multifaceted value of rapidly identifying chemokine and/or cytokine biomarkers in future pandemic settings. Indeed, there has to date been negligible incorporation of immune signals into diagnostic algorithms that guide clinical decision-making. Rather, current point-of-care tests rely mainly on biochemical markers including acute phase proteins released several hours after sentinel innate host - pathogen interactions and which do not directly mirror specific processes underpinning downstream patient deterioration.

The value of prognostic tests in emerging infections, when healthcare systems can be severely strained, is illustrated by the very many candidate approaches deployed in COVID-19, some with acceptable performance^2^. Nonetheless, optimal test suitability is dictated by many factors, reflecting which we have sought a parsimonious platform broadly applicable wherever clinical laboratory testing operates, and reliant only on patient sera. Adding IP10 measurement alone to clinical laboratory blood tests performed comparably to adding measurements of several cytokines and chemokines and is obviously cheaper and more straightforward, particularly given the development of a clinical assay that was employed in this study. Additionally, the IP10 data can be simply integrated into other clinical laboratory tests, obviating the need for a busy clinician’s time and input to calculate a score. Also, and unlike NEWS2, there is no reliance on physiological parameters manually recorded on paper or on e-noting systems, both of which are commonly unlinked to laboratory test output software.

Adding IP10 to clinical laboratory tests performed better than adding IL-6, high levels of which have been directly implicated in contributing to pathology, efficaciously treated in some cases by tocilizumab^34^. Possibly this is because the greater relevance of IL-6 is contemporaneous with clinical deterioration, whereas high IP10 levels anticipate deterioration at earlier timepoints. Thus, one can ask which aspects of IP10 biology might explain its prognostic capacity in combination with other tests?

Clearly, IP10 is not a direct and obligatory determinant of COVID-19 pathology because its levels were not ostensibly high in vaccinated patients in India progressing to ICU. In other settings, however, IP10 levels mirror SARS-CoV-2 viral load in the nasopharynx during acute infection^38^, whilst sustained elevation is detectable up to 4 months post infection in long COVID-19, consistent with hypotheses that viral persistence drives a subset of symptoms and likewise consistent with data showing that up to 1 in 200 patients have detectable SARS-CoV-2 RNA for at least two months post-infection^39^. This linkage to viraemia might reflect IP10 upregulation by Type I IFNs responding to virus. *A priori* this might seem paradoxical given widely reported settings of severe disease associated with Type I IFN deficiency^40–42^. However, a second and conceivably major aspect of IP10 biology is that its expression by monocytes is at least as well induced by Type II IFN (IFNγ)^43^. in which regard it has been used as a facile, quantitative biomarker for T cell activation^44^.

We therefore hypothesised that elevated IP10 levels at hospital admission most probably reflect T cell hyperactivation that may portend T cytopenia attributable to activation-induced cell death and/or other causes. This hypothesis received support from our unbiased, high-content immune-profiling showing that cellular immune correlates of high IP10 levels and severity were overtly enriched in T cells and activation markers seemingly followed rapidly by cytopenia. Moreover, these observations held across two very different COVID-19 cohorts in London and Bangalore, respectively, and from very different times in the evolution of the virus and treatment regimens including immunosuppression. This consistency, which was not shown by other correlates of severity, underlines the likelihood that severe COVID-19 is caused by gross immunological dysregulation, e.g., exaggerated myeloid cell activity, that is downstream of early T cell dysfunction and loss. Why T cells are overtly hyperactivated in some patients and not others remains to be determined, but might in part reflect failure to limit virus spread^45^. Irrespective of cause, it seems highly possible that interventions limiting T cell activation and loss, might be of clinical benefit.

One such intervention may be vaccination itself. In the highly heterogeneous Bangalore cohort, experiencing a healthcare system under severe strain across 2022, IP10 levels showed valuable high sensitivity in prognosticating those who would terminally deteriorate in the ICU. By contrast, IP10 levels did not discriminate disease severity for other patients the great majority of whom were vaccinated. We hypothesise that this reflects the regulated production of Type II IFN by antigen-specific T cells making textbook responses to infection^46^, that include scheduled development of neutralizing antibodies^47^. Thus, the prognostic value of IP10 levels might be primarily in unvaccinated persons who most often will be the great majority at the beginning of a pandemic. This in turn emphasizes how biomarkers need to be applied according to the status of a pandemic within society.

Possibly, IP10 is of particular relevance to the immune dysregulation provoked by coronavirus infection, in which regard it seems noteworthy that an IP10 processing enzyme, DPP4, is a receptor for MERS virus^48^. Nonetheless, we believe that our data provide the rationale to rapidly evaluate the potential of bespoke, disease-relevant, easily measurable immune markers including IP10 as diagnostic adjuncts to assess severity or predict deterioration in newly emerging infections. This seems particularly appropriate as reference ranges for different cytokines and chemokines are being developed based on the analysis of tens of thousands of healthy controls. By contrast, whereas re-purposing of already validated and familiar scores including NEWS2 might seem an attractive approach for rapid evaluation of patient prognosis in novel human contagions, an inherent disadvantage with NEWS2 is its reliance on physiological parameters that mostly bear imprecise relationships to any one disease process. Those limitations are highlighted by the PRIEST trial of 20,891 COVID-19 patients recruited from the Emergency Department in 2020, which demonstrated poor specificity (0.28) at a NEWS2 threshold of >1, albeit that the sensitivity was high (0.96)^26^. This would mean that while few at-risk patients would be missed, the “false-positives” could place a huge burden on treatment resource, compounding requirement for manual entry into a risk calculator: indeed, additional time is a known barrier for implementation of prognostic scores in general, irrespective of their accuracy^49^.

### Limitations of the study

As for all models, the data generated are suitable for the derivation cohort, and performance is likely to vary as cohorts and treatments change. Furthermore, for estimating the true value provided by the models a full cost-effectiveness analysis quantifying all benefits and harms of model implementation should be performed instead of simple decision curves. While model performance characteristics were acceptable in the UK validation cohort (2021), the models that could be tested in India in 2022 performed relatively poorly when trying to predict detoriation in terms of ICU admission or death. Nevertheless, although based on a small number of deaths observed only among individuals without a recorded vaccination status or who were known to be unvaccinated, the linkage between IP10 and death remained preserved in this more recent cohort of patients from India.

Finally, our study has emphasised the capacity of chemokine / cytokine measurements to connect to probable causes of pathogenesis *via* the implementation of high-content, high-throughput immunophenotyping, i.e., from >1800 cell types and states measured, unbiased associations with IP10 levels could be filtered to those also associated with severity in two entirely different settings. In summarizing these statistical associations, we filtered the parameters to report single instances of association between a given phenotypic marker (+ or −) with a given cell type, thereby avoiding double counting and hence biased interpretation. Nonetheless, we acknowledge that this may reduce the resolution of the data, with potential to occasionally miss some relevant associations. Further refinement of these methods is ongoing.

## Data Availability

All data produced in the present study are available upon reasonable request to the authors

## Acknowledgments

We thank Dr. Francesca di Rosa and Dr. Irene del Molino del Barrio, for advice, guidance and assistance at various stages of the project.This research was supported by a Medical Research Council grant, CARDINNATE. AD was supported by an Academy of Medical Sciences Starter Grant for Clinical Lecturers (SGL019\1004) and a National Institute for Health Research (NIHR) Academic Clinical Lectureship. LBS is funded by the Medical Research Council (MR/W025140/1). KBP is supported by the Medical Research Foundation (MRF_160-0017-ELP-POUW-C0909) and NIHR Health Protection Research Unit (HPRU) in Healthcare Associated Infections and Antimicrobial Resistance at the University of Oxford in partnership with UK Health Security Agency (UKHSA; NIHR200915). The views expressed are those of the authors and not necessarily those of the NIHR, UKHSA, NHS, or Department of Health.

## Declaration of interests

DM, GD, BT, TH are fulltime employees for IMU biosciences. ACH is a consultant for Takeda Pharmaceuticals, Prokarium, TransImmune, AG, and ImmunoQure, AH, and receives research funds from Takeda Pharmaceuticals, but the work described here is outside of those interests and none of the named parties conceived of, undertook, influenced on commented on the contents of this study. All other authors declare no competing interests.

---

**Supplementary Table 1.** Demographics of UK Discovery and Validation cohorts. P values were compared using chi-squared for categorical variables or by two-tailed Mann Whitney U-test for non-categorical variables. Where appropriate, n numbers for missing data are shown.

| Variables | Discovery | Validation | *P* value |
| --- | --- | --- | --- |
| N | 278 | 176 |  |
| Age [median (Q1-Q3)] | 59 (48-73.3) | 60 (48-75) | 0.5 |
| BMI [median (Q1-Q3)] | 25.6 (21.7-30.3) | 26.9 (23-31.2) | 0.073 |
| BMI [missing N] | 25 | 55 |  |
| Male | 182[65.5%] | 106[60.2%] | 0.26 |
| Hypertension | 110[39.6%] | 66[37.5%] | 0.66 |
| Diabetes | 90[32.4%] | 41[23.3%] | 0.038 |
| Chronic lung disease [non asthma] | 29[10.4%] | 22[12.5%] | 0.497 |
| Asthma | 23[8.3%] | 23[13.1%] | 0.099 |
| Ischaemic heart disease | 20[7.2%] | 25[14.2%] | 0.015 |
| Congestive cardiac failure | 11[4%] | 7[4%] | 0.99 |
| Hyperlipidaemia | 30[10.8%] | 26[14.8%] | 0.21 |
| Malignancy | 23[8.3%] | 17[9.7%] | 0.61 |
| Ethnicity |  |  |  |
| Asian | 31 [12.2%] | 25 [15.8%] | 0.297 |
| Black | 94 [37%] | 43 [27.2%] | 0.04 |
| Caucasian | 123 [48.4%] | 80 [50.6%] | 0.66 |
| Latin | 3 [1.2%] | 7 [4.4%] | 0.037 |
| Other | 3 [1.2%] | 3 [1.9%] | 0.55 |
| Ethnicity [missing N] | 24 | 18 |  |

**Supplementary Table 2.** Missing data in Discovery and Validation cohorts.

|  | Discovery | Validation | All | % Missing (All) |
| --- | --- | --- | --- | --- |
| N | 278 | 176 | 454 |  |
| N Missing(IL1b) | 0 | 0 | 0 | 0 |
| N Missing(TNFa) | 0 | 0 | 0 | 0 |
| N Missing(IP10) | 0 | 0 | 0 | 0 |
| N Missing(IL6) | 0 | 0 | 0 | 0 |
| N Missing(IL10) | 0 | 0 | 0 | 0 |
| N Missing(IL8) | 0 | 0 | 0 | 0 |
| N Missing(IL12p70) | 0 | 0 | 0 | 0 |
| N Missing(IFNa2) | 0 | 0 | 0 | 0 |
| N Missing(GMCSF) | 0 | 0 | 0 | 0 |
| N Missing(Neutrophils) | 1 | 0 | 1 | 0.2 |
| N Missing(Lymphocytes) | 1 | 0 | 1 | 0.2 |
| N Missing(Platelets) | 1 | 0 | 1 | 0.2 |
| N Missing(IFNb) | 1 | 0 | 1 | 0.2 |
| N Missing(Creatinine) | 3 | 0 | 3 | 0.7 |
| N Missing(CRP) | 6 | 5 | 11 | 2.4 |
| N Missing(Albumin) | 20 | 21 | 41 | 9.0 |
| **Excluded** |  |  |  |  |
| N Missing(Bilirubin) | 22 | 24 | 46 | 10.1 |
| N Missing(Glucose) | 30 | 21 | 51 | 11.2 |
| N Missing(Alanine Aminotransferase) | 40 | 29 | 69 | 15.2 |
| N Missing(Ferritin) | 96 | 87 | 183 | 40.3 |
| N Missing(Urea) | 134 | 61 | 195 | 43.0 |
| N Missing(D-dimer) | 139 | 97 | 236 | 52.0 |
| N Missing(Troponin) | 150 | 87 | 237 | 52.2 |

**Supplementary Table 3.** Demographics of India cohorts shown in Figure 4. P values were compared using chi-squared for categorical variables or by two-tailed Kruskal-Wallis with Dunn’s post hoc correction for age.

|  | India non-COVID-19<br>(a) | India COVID-19 survived<br>(b) | India COVID-19 died<br>(c) | P value |
| --- | --- | --- | --- | --- |
| N | 100 | 79 | 9 |  |
| Age [median (Q1-Q3)] | 35 (26-49) | 49 (35-61) | 65 (53-75) | a vs b P=0.0006; b vs c<br>P=0.044; a vs c<br>P=0.0001 |
| Male | 55 (55%) | 40 (50.6%) | 3 (33.3%) | 0.43 |
| Hypertension | 13 (13%) | 34 (43%) | 4 (44.4%) | 0.00002 |
| Diabetes | 14 (14%) | 29 (36.7%) | 4 (44.4%) | 0.00089 |
| Chronic Lung Disease | 10 (10%) | 16 (20.3%) | 3 (33.3%) | 0.05 |
| Coronary Artery Disease | 2 (2%) | 9 (11.4%) | 7 (77.8%) | 9.9 x 10<sup>-13</sup> |
| Hyperlipidaemia | 3 (3%) | 6 (7.6%) | 0 (0%) | 0.28 |
| Malignancy | 1 (1%) | 5 (6.3%) | 2 (22.2%) | 0.005 |

**Supplementary Table 4.**
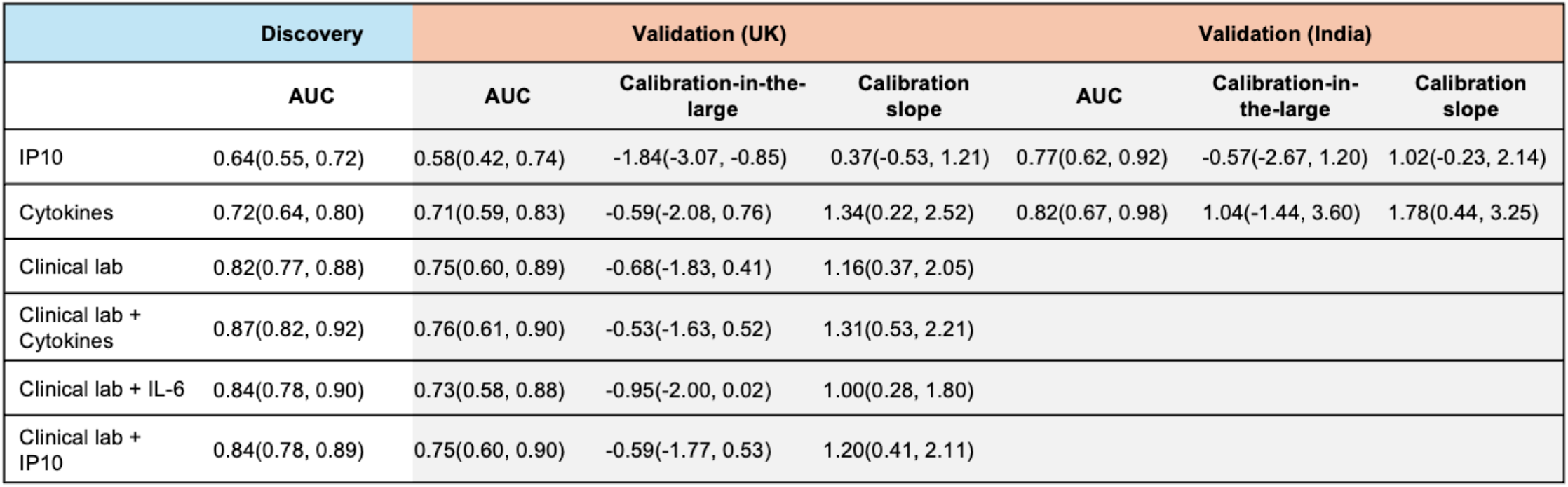
Sensitivity analyses for all cohorts for mortality only.

| Discovery |  | Validation (UK) |  |  | Validation (India) |  |  |
| --- | --- | --- | --- | --- | --- | --- | --- |
|  | AUC | AUC | Calibration-in-the-large | Calibration slope | AUC | Calibration-in-the-large | Calibration slope |
| IP10 | 0.64(0.55, 0.72) | 0.58(0.42, 0.74) | -1.84(-3.07, -0.85) | 0.37(-0.53, 1.21) | 0.77(0.62, 0.92) | -0.57(-2.67, 1.20) | 1.02(-0.23, 2.14) |
| Cytokines | 0.72(0.64, 0.80) | 0.71(0.59, 0.83) | -0.59(-2.08, 0.76) | 1.34(0.22, 2.52) | 0.82(0.67, 0.98) | 1.04(-1.44, 3.60) | 1.78(0.44, 3.25) |
| Clinical lab | 0.82(0.77, 0.88) | 0.75(0.60, 0.89) | -0.68(-1.83, 0.41) | 1.16(0.37, 2.05) |  |  |  |
| Clinical lab + Cytokines | 0.87(0.82, 0.92) | 0.76(0.61, 0.90) | -0.53(-1.63, 0.52) | 1.31(0.53, 2.21) |  |  |  |
| Clinical lab + IL-6 | 0.84(0.78, 0.90) | 0.73(0.58, 0.88) | -0.95(-2.00, 0.02) | 1.00(0.28, 1.80) |  |  |  |
| Clinical lab + IP10 | 0.84(0.78, 0.89) | 0.75(0.60, 0.90) | -0.59(-1.77, 0.53) | 1.20(0.41, 2.11) |  |  |  |

**Supplementary Table 5.**
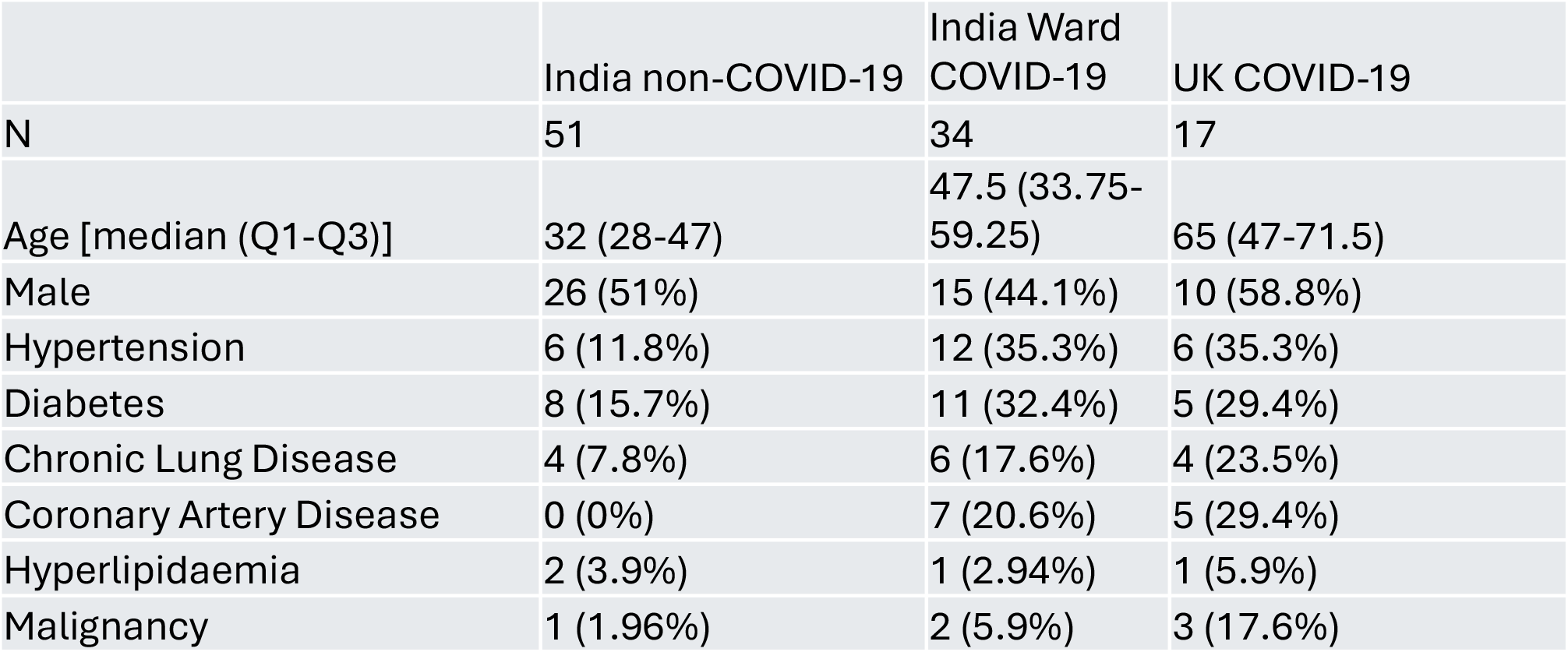
Immunophenotyping cohort characteristics. Linked to figures 5-7.

**Supplementary Table 6.** Antibody panels (see STAR Methods)

| T panel |  | APC panel |  |
| --- | --- | --- | --- |
| Marker | Clone | Marker | Clone |
| Live Dead Blue | - | Live Dead Blue | - |
| CD14 | HCD14 | CD14 | HCD14 |
| CD16 | 3G8 | CD16 | 3G8 |
| CD19 | HIB19 | CD19 | HIB19 |
| CD27 | OA17A18 | CD27 | OA17A18 |
| CD279/PD-1 | EH12.1 | CD279/PD-1 | EH12.1 |
| CD3 | OKT3 | CD3 | OKT3 |
| CD38 | HB7 | CD38 | HB7 |
| CD4 | SK3 | CD4 | SK3 |
| CD45 | H130 | CD45 | H130 |
| CD5 | UCHT2 | CD5 | UCHT2 |
| CD56 | 5.1H11 | CD56 | 5.1H11 |
| CD8 | QA18A37 | CD8 | QA18A37 |
| HLA-DR | L243 | HLA-DR | L243 |
| TCRgd | 11F2 | TCRgd | 11F2 |
| CCR10 | 1B5 | CD10 | HI10a |
| CD103/ITGAE | Ber-ACT8 | CD11c | B-ly6 |
| CD122/IL2RB | Mik-β3 | CD123 | 7G3 |
| CD127 | HIL-7R-M21 | CD138 | MI15 |
| CD161/KLRB1 | HP-3G10 | CD141 | 1A4 |
| CD183/CXCR3 | 1C6/CXCR3 | CD15 | HI98 |
| CD185/CXCR5 | RF8B2 | CD1c | L161 |
| CD194/CCR4 | 1G1 | CD24 | ML5 |
| CD196/CCR6 | G034E3 | CD267/TACI | 1A1 |
| CD197/CCR7 | 2-L1-A | CD274/PD-L1 | MIH1 |
| CD223/LAG-3 | T47-530 | CD294/CRTH2 | BM16 |
| CD25 | CD25-3G10 | CD303 | 201A |
| CD278/ICOS | DX29 | CD319 | 162.1 |
| CD28 | CD28.2 | CD335/NKp46 | 9E2/NKp46 |
| CD31 | WM59 | CD337/NKp30 | p30-15 |
| CD366/TIM-3 | 7D3 | CD40 | 5C3 |
| CD39 | TU66 | CD43 | 1G10 |
| CD45RA | HI100 | CD62L | DREG-56 |
| CD45RO | UCHL1 | CD69 | FN50 |
| CD57 | HNK-1 | CD86 | 2331 (FUN-1) |
| CD95 | DX2 | IgD | IA6-2 |
| KLRG1 | SA231A2 | IgM | G20-127 |
| TCR Va24-Ja18 | 6B11 |  |  |
| TCR Va7.2 | REA179 |  |  |
| TCR Vδ1 | REA173 |  |  |
| TCR Vδ2 | REA771 |  |  |
| TIGIT | 741182 |  |  |

**Supplementary Figure 1.**
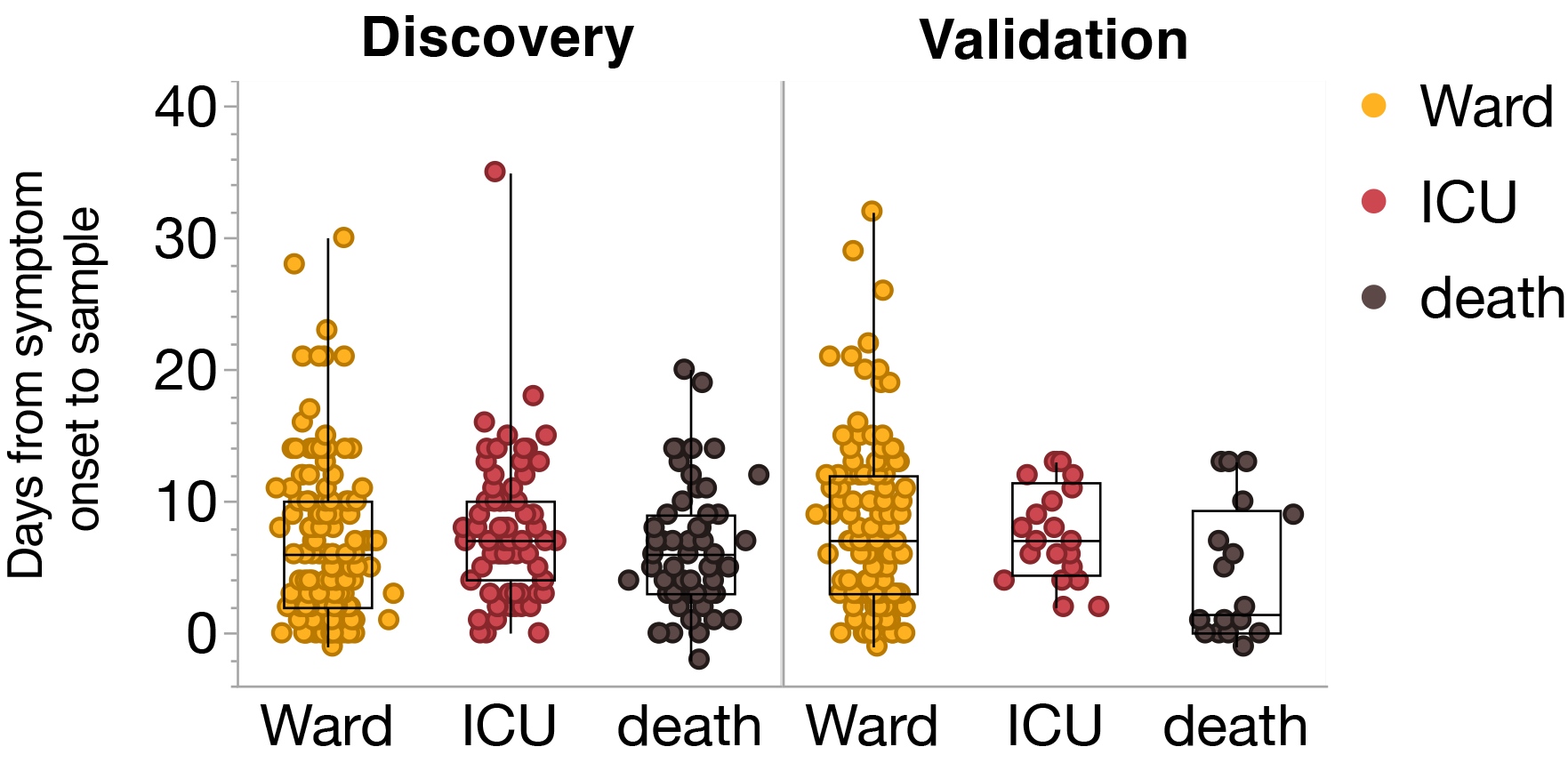
Time from symptom onset to sampling. The number of days between symptom onset to sample. Box plots; central line denotes the median, upper and lower lines of box represent the 75th and 25th percentiles respectively and whiskers show the range (minimum to maximum value).

**Supplementary Figure 2.**
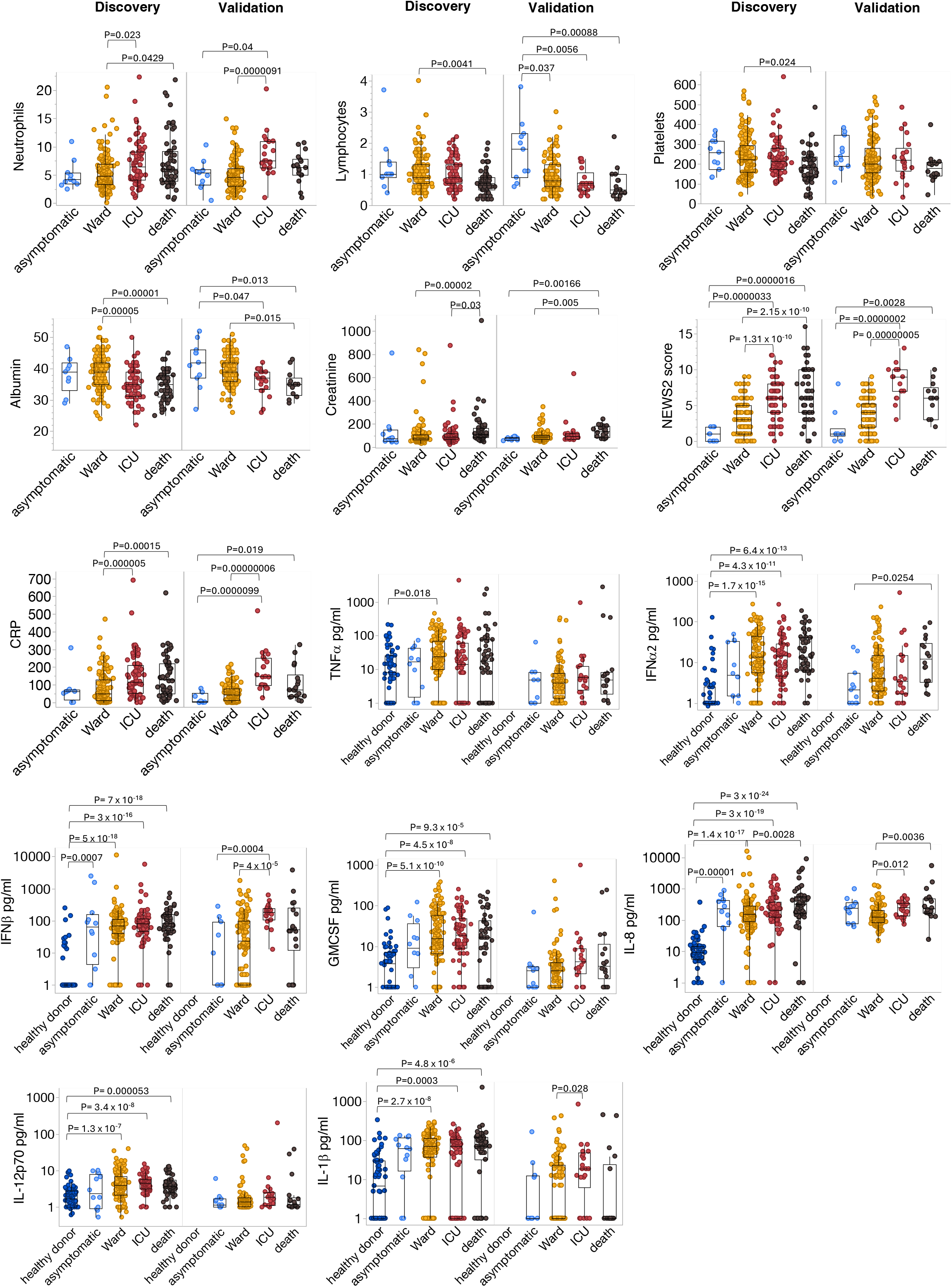
Admission laboratory analytes, cytokines and NEWS2 score. Linked to Figure 2. Comparison of six clinical laboratory analytes (those with <10% missing values), seven cytokines and NEWS2 score between patient groups (measured at hospital admission) in Discovery and Validation cohorts. *P*-values were generated by a Kruskal–Wallis test with Dunn’s post hoc correction. Box plots show median, first quartile (Q1) and 3^rd^ quartile (Q3) and whiskers extend between Q1-1.5 x IQR (interquartile range) and Q3+1.5 x IQR.

**Supplementary Figure 3.**
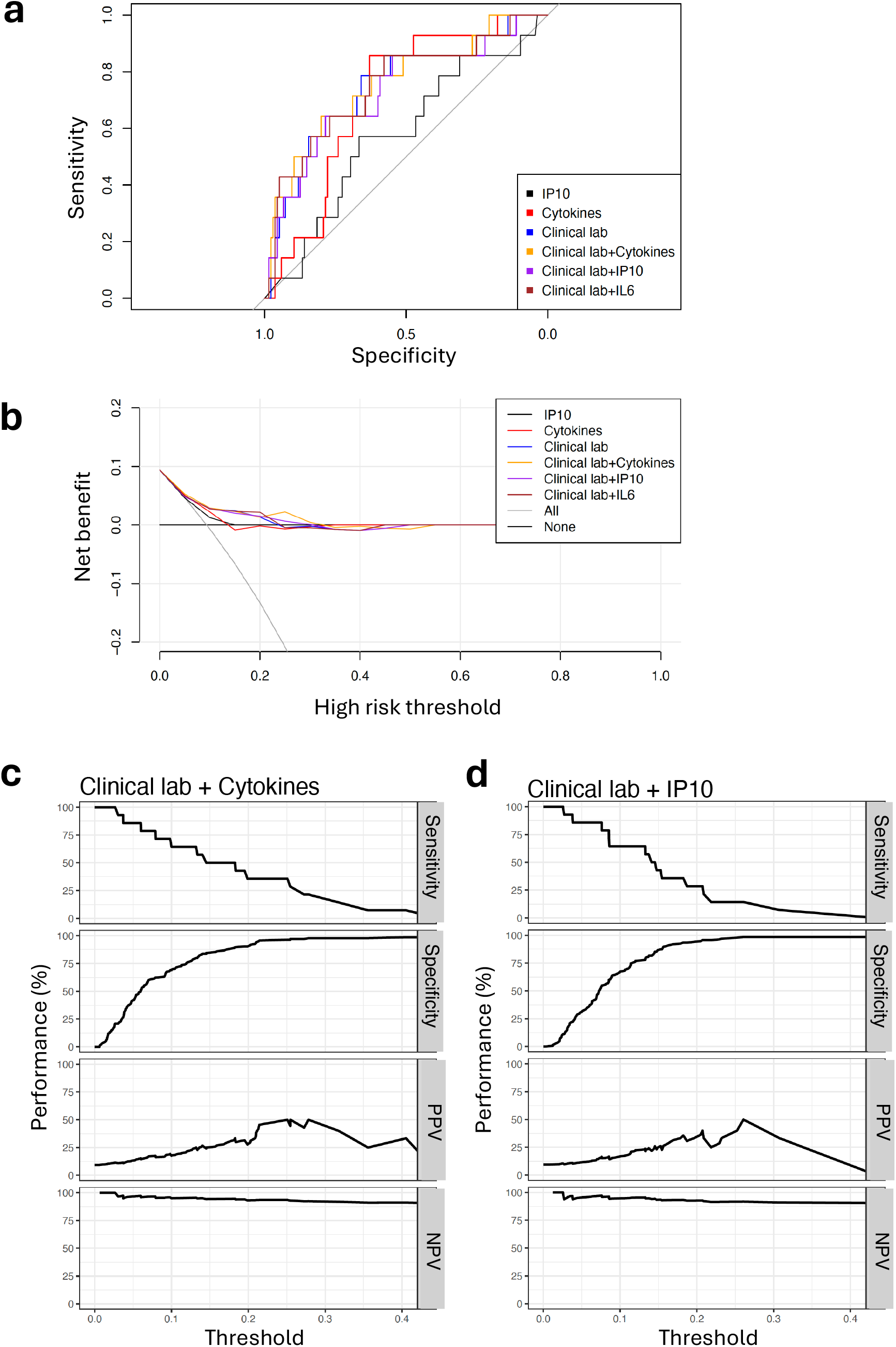
Sensitivity analysis for mortality alone in the UK Validation cohort. a, Comparison of area under the receiver operating characteristic curve (AUROC) among six models in the validation cohort. Models are recalibrated to the validation data. b, Decision curve analysis across eight models in the validation cohort. Net benefit is plotted for each model compared with the treat-all and treat-none approaches. Models are recalibrated to the validation data. Sensitivity, specificity, positive predictive values (PPVs) and negative predictive values (NPVs) of the c, clinical lab and cytokines or d, clinical lab and IP10 model in the validation cohort, according to the full range of probability thresholds for the model prediction.

**Supplementary Figure 4.**
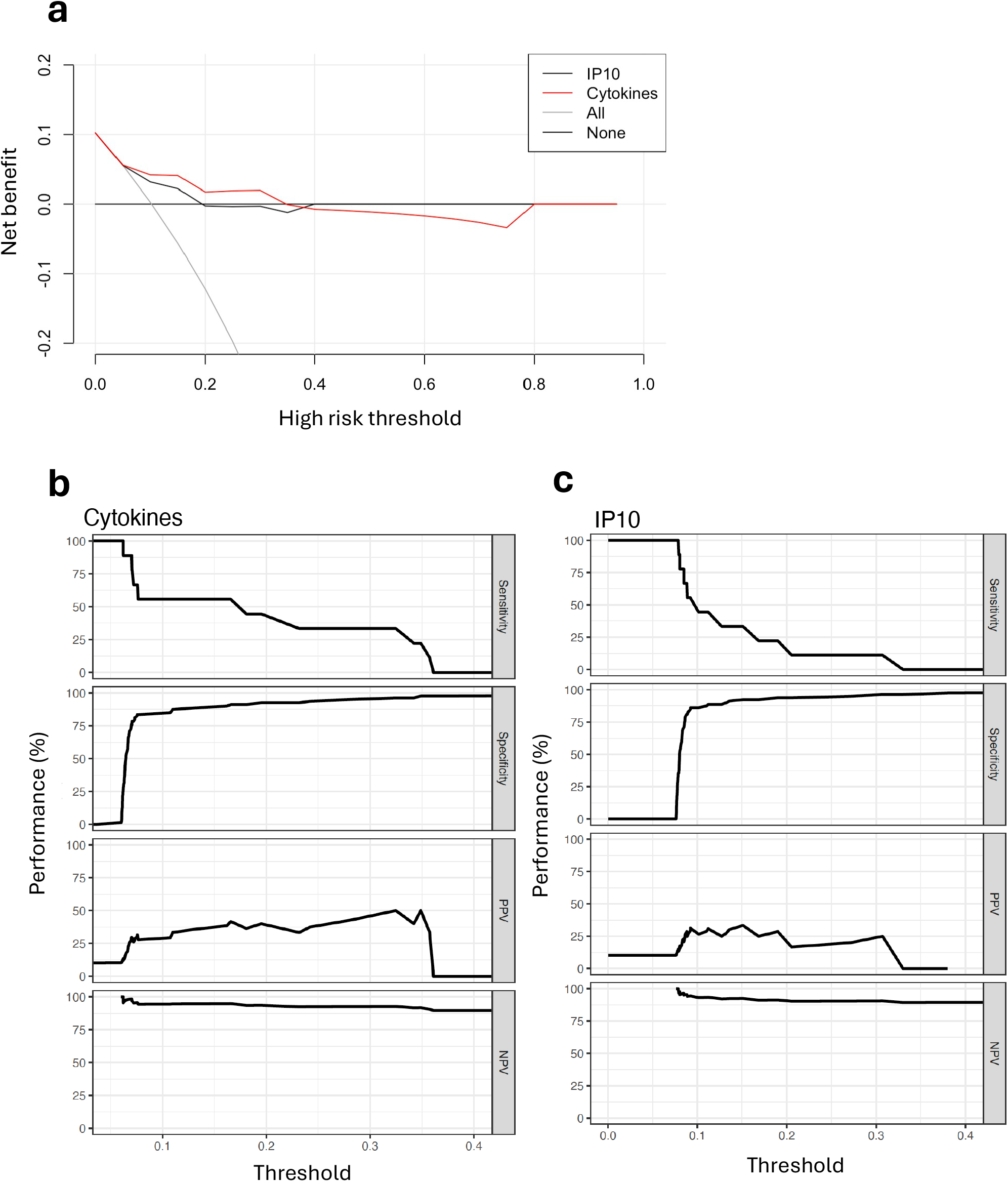
Sensitivity analysis for mortality alone in the India Validation cohort. a, Decision curve analysis across four models in the India Validation cohort. Net benefit is plotted for each model compared with the treat-all and treat-none approaches. Models are recalibrated to the validation data. Sensitivity, specificity, positive predictive values (PPVs) and negative predictive values (NPVs) of b, cytokines or c, IP10 models in the validation cohort, according to the full range of probability thresholds for the model prediction.

## Notes

### Funding Statement

This research was funded by a Medical Research
Council grant CARDINNATE. AD was supported by an Academy of Medical Sciences Starter
Grant for Clinical Lecturers (SGL019\1004) and a National Institute for Health Research (NIHR)
Academic Clinical Lectureship. LBS is funded by the Medical Research Council
(MR/W025140/1). KBP is supported by the Medical Research Foundation (MRF_160-0017-
ELP-POUW-C0909) and NIHR Health Protection Research Unit (HPRU) in Healthcare
Associated Infections and Antimicrobial Resistance at the University of Oxford in partnership
with UK Health Security Agency (UKHSA NIHR200915).

### Author Declarations

The South Central Hampshire B Research Ethics committee gave ethical approval for this work The Institutional Ethics Committee at St. Johns Medical College Hospital Bangalore gave ethical approval for this work

## References

1. Coronavirus (COVID-19) action plan. GOV.UK https://www.gov.uk/government/publications/coronavirus-action-plan.

2. Wynants, L. et al. Prediction models for diagnosis and prognosis of covid-19: systematic review and critical appraisal. BMJ 369, m1328 (2020).

3. Riley, R. D. et al. A guide to systematic review and meta-analysis of prognostic factor studies. BMJ 364, k4597 (2019).

4. Laing, A. G. et al. A dynamic COVID-19 immune signature includes associations with poor prognosis. Nat Med 26, 1623–1635 (2020).

5. Lucas, C. et al. Longitudinal analyses reveal immunological misfiring in severe COVID-19. Nature 584, 463–469 (2020).

6. Thwaites, R. S., et al. Inflammatory profiles across the spectrum of disease reveal a distinct role for GM-CSF in severe COVID-19. Sci Immunol 6, eabg9873 (2021).

7. Mathew, D. et al. Deep immune profiling of COVID-19 patients reveals distinct immunotypes with therapeutic implications. Science 369, eabc8511 (2020).

8. Kuri-Cervantes, L. et al. Comprehensive mapping of immune perturbations associated with severe COVID-19. Science Immunology 5, eabd7114 (2020).

9. Gupta, R. K. et al. Development and validation of the ISARIC 4C Deterioration model for adults hospitalised with COVID-19: a prospective cohort study. The Lancet Respiratory Medicine 9, 349– 359 (2021).

10. Knight, S. R. et al. Risk stratification of patients admitted to hospital with covid-19 using the ISARIC WHO Clinical Characterisation Protocol: development and validation of the 4C Mortality Score. BMJ 370, m3339 (2020).

11. Knight, S. R. et al. Prospective validation of the 4C prognostic models for adults hospitalised with COVID-19 using the ISARIC WHO Clinical Characterisation Protocol. Thorax 77, 606–615 (2022).

12. Kartsonaki, C. et al. Characteristics and outcomes of an international cohort of 600 000 hospitalized patients with COVID-19. Int J Epidemiol 52, 355–376 (2023).

13. Starke, K. R. et al. The isolated effect of age on the risk of COVID-19 severe outcomes: a systematic review with meta-analysis. BMJ Global Health 6, e006434 (2021).

14. Tay, J. K. & Tibshirani, R. Reluctant Generalised Additive Modelling. Int Stat Rev 88, S205– S224 (2020).

15. Hastie, T. & Tibshirani, R. Generalized Additive Models. Statistical Science 1, 297–310 (1986).

16. Yu, G., Bien, J. & Tibshirani, R. Reluctant Interaction Modeling. Preprint at 10.48550/arXiv.1907.08414 (2023).

17. Steyerberg, E. W. & Vergouwe, Y. Towards better clinical prediction models: Seven steps for development and an ABCD for validation. European Heart Journal vol. 35 1925–1931 Preprint at 10.1093/eurheartj/ehu207 (2014).

18. Toll, D. B., Janssen, K. J. M., Vergouwe, Y. & Moons, K. G. M. Validation, updating and impact of clinical prediction rules: A review. Journal of Clinical Epidemiology vol. 61 1085–1094 Preprint at 10.1016/j.jclinepi.2008.04.008 (2008).

19. Steyerberg, E. W. Clinical Prediction Models. (Springer New York, New York, NY, 2009). doi:10.1007/978-0-387-77244-8.

20. Vickers, A. J., van Calster, B. & Steyerberg, E. W. A simple, step-by-step guide to interpreting decision curve analysis. Diagnostic and Prognostic Research 3, 1–8 (2019).

21. Gu, Z., Gu, L., Eils, R., Schlesner, M. & Brors, B. circlize implements and enhances circular visualization in R. Bioinformatics 30, 2811–2812 (2014).

22. Turtle, L. et al. Changes in hospital mortality in patients with cancer during the COVID-19 pandemic (ISARIC-CCP-UK): a prospective, multicentre cohort study. The Lancet Oncology 25, 636–648 (2024).

23. Huang, C. et al. Clinical features of patients infected with 2019 novel coronavirus in Wuhan, China. Lancet 395, 497–506 (2020).

24. Wang, D. et al. Clinical Characteristics of 138 Hospitalized Patients With 2019 Novel Coronavirus-Infected Pneumonia in Wuhan, China. JAMA 323, 1061–1069 (2020).

25. Gao, M. et al. Associations between body-mass index and COVID-19 severity in 6·9 million people in England: a prospective, community-based, cohort study. The Lancet Diabetes & Endocrinology 9, 350–359 (2021).

26. Thomas, B. et al. Prognostic accuracy of emergency department triage tools for adults with suspected COVID-19: the PRIEST observational cohort study. Emerg Med J 38, 587–593 (2021).

27. Docherty, A. B. et al. Features of 20 133 UK patients in hospital with covid-19 using the ISARIC WHO Clinical Characterisation Protocol: prospective observational cohort study. BMJ 369, m1985 (2020).

28. National Early Warning Score (NEWS) 2. https://www.rcp.ac.uk/improving-care/resources/national-early-warning-score-news-2/.

29. Lorè, N. I. et al. CXCL10 levels at hospital admission predict COVID-19 outcome: hierarchical assessment of 53 putative inflammatory biomarkers in an observational study. Molecular Medicine 27, 129 (2021).

30. Blot, M. et al. CXCL10 could drive longer duration of mechanical ventilation during COVID-19 ARDS. Crit Care 24, 632 (2020).

31. Samaras, C. et al. Interferon gamma-induced protein 10 (IP-10) for the early prognosis of the risk for severe respiratory failure and death in COVID-19 pneumonia. Cytokine 162, 156111 (2023).

32. Trigo-Rodríguez, M. et al. Role of IP-10 to Predict Clinical Progression and Response to IL-6 Blockade With Sarilumab in Early COVID-19 Pneumonia. A Subanalysis of the SARICOR Clinical Trial. Open Forum Infectious Diseases 10, ofad133 (2023).

33. Lev, S. et al. Observational cohort study of IP-10’s potential as a biomarker to aid in inflammation regulation within a clinical decision support protocol for patients with severe COVID-19. PLoS One 16, e0245296 (2021).

34. Abani, O. et al. Tocilizumab in patients admitted to hospital with COVID-19 (RECOVERY): a randomised, controlled, open-label, platform trial. The Lancet 397, 1637–1645 (2021).

35. The REMAP-CAP Investigators. Interleukin-6 Receptor Antagonists in Critically Ill Patients with Covid-19. New England Journal of Medicine 384, 1491–1502 (2021).

36. Diao, B. et al. Reduction and Functional Exhaustion of T Cells in Patients With Coronavirus Disease 2019 (COVID-19). Front. Immunol. 11, (2020).

37. Chen, G. et al. Clinical and immunological features of severe and moderate coronavirus disease 2019. J Clin Invest 130, 2620–2629 (2020).

38. Cheemarla, N. R. et al. Dynamic innate immune response determines susceptibility to SARS-CoV-2 infection and early replication kinetics. J Exp Med 218, e20210583 (2021).

39. Ghafari, M. et al. Prevalence of persistent SARS-CoV-2 in a large community surveillance study. Nature 626, 1094–1101 (2024).

40. McNab, F., Mayer-Barber, K., Sher, A., Wack, A. & O’Garra, A. Type I interferons in infectious disease. Nat Rev Immunol 15, 87–103 (2015).

41. Bastard, P. et al. Preexisting autoantibodies to type I IFNs underlie critical COVID-19 pneumonia in patients with APS-1. J Exp Med 218, e20210554 (2021).

42. Bastard, P. et al. Autoantibodies against type I IFNs in patients with life-threatening COVID-19. Science 370, eabd4585 (2020).

43. Luster, A. D., Unkeless, J. C. & Ravetch, J. V. Gamma-interferon transcriptionally regulates an early-response gene containing homology to platelet proteins. Nature 315, 672–676 (1985).

44. Schwarz, M. et al. Rapid, scalable assessment of SARS-CoV-2 cellular immunity by whole-blood PCR. Nat Biotechnol 40, 1680–1689 (2022).

45. Moss, P. The T cell immune response against SARS-CoV-2. Nat Immunol 23, 186–193 (2022).

46. Bertoletti, A., Le Bert, N. & Tan, A. T. SARS-CoV-2-specific T cells in the changing landscape of the COVID-19 pandemic. Immunity 55, 1764–1778 (2022).

47. Lucas, C. et al. Delayed production of neutralizing antibodies correlates with fatal COVID-19. Nat Med 27, 1178–1186 (2021).

48. Casrouge, A. et al. Discrimination of agonist and antagonist forms of CXCL10 in biological samples. Clin Exp Immunol 167, 137–148 (2012).

49. Sharma, V. et al. Adoption of clinical risk prediction tools is limited by a lack of integration with electronic health records. BMJ Health Care Inform 28, e100253 (2021).

